# Estimating salt consumption in 49 low- and middle-income countries: Development, validation and application of a machine learning model

**DOI:** 10.1101/2021.08.31.21262944

**Authors:** Wilmer Cristobal Guzman-Vilca, Manuel Castillo-Cara, Rodrigo M. Carrillo-Larco

**Affiliations:** School of Medicine “Alberto Hurtado”, Universidad Peruana Cayetano Heredia, Lima, Peru; CRONICAS Centre of Excellence in Chronic Diseases, Universidad Peruana Cayetano Heredia, Lima, Peru; Sociedad Científica de Estudiantes de Medicina Cayetano Heredia (SOCEMCH), Universidad Peruana Cayetano Heredia, Lima, Peru; Universidad de Lima, Lima, Peru.; Department of Epidemiology and Biostatistics, School of Public Health, Imperial College London, London, UK

**Keywords:** artificial intelligence, deep learning, cardio-metabolic risk factors, cardiovascular health, global health, population health

## Abstract

**Background:** Global targets to reduce salt intake have been proposed but their monitoring is challenged by the lack of population-based data on salt consumption. We developed a machine learning (ML) model to predict salt consumption based on simple predictors, and applied this model to national surveys in low- and middle-income countries (LMICs).

**Methods:** Pooled analysis of WHO STEPS surveys. We used 19 surveys with spot urine samples for the ML model derivation and validation; we developed a supervised ML regression model based on: sex, age, weight, height, systolic and diastolic blood pressure. We applied the ML model to 49 new STEPS surveys to quantify the mean salt consumption in the population.

**Results:** The pooled dataset in which we developed the ML model included 45,152 people. Overall, there were no substantial differences between the observed (8.1 g/day (95% CI: 8.0-8.2 g/day)) and ML-predicted (8.1 g/day (95% CI: 8.1-8.2 g/day)) mean salt intake (p= 0.065). The pooled dataset where we applied the ML model included 157,699 people; the overall predicted mean salt consumption was 8.1 g/day (95% CI: 8.1-8.2 g/day). The countries with the highest predicted mean salt intake were in Western Pacific. The lowest predicted intake was found in Africa. The country-specific predicted mean salt intake was within reasonable difference from the best available evidence.

**Conclusions:** A ML model based on readily available predictors estimated daily salt consumption with good accuracy. This model could be used to predict mean salt consumption in the general population where urine samples are not available.

**Funding:** Wellcome Trust (214185/Z/18/Z)

## INTRODUCTION

The association between high sodium/salt intake and high blood pressure, a major risk factor of cardiovascular diseases (CVD), is well-established.^1–3^ More than 1.7 million CVD deaths were attributed to a diet high in sodium in 2019, with ∼90% of these deaths occurring in low- and middle-income countries (LMICs).^4, 5^ Consequently, salt reduction has been included in international goals: the World Health Organization (WHO) recommendation of limiting salt consumption to <5 g/day,^2^ and the agreement by the WHO state members of a 30% relative reduction in mean population salt intake by 2025.^6^ Because available evidence suggests that sodium/salt consumption is higher than the global targets,^7–9^ we need timely and consistent data of sodium/salt consumption in the general population to track progress of salt reduction targets.

Global efforts have been made to produce comparable estimates of sodium/salt intake for all countries.^7^ Similarly, researchers have summarized all the available evidence in specific world regions.^8, 9^ Although the global endeavor was based on the gold standard method to assess sodium/salt intake (i.e., 24-hour urine sample), their estimates were up to 2010.^7^ Therefore, robust and comparable sodium/salt intake estimates for all countries lack for the last ten years. The regional endeavors summarized population-based evidence, yet they conducted study-level meta-analyses in which the original studies could have followed different laboratory methods, and they did not study all countries in the region. Therefore, comparability across studies could be limited and evidence lacks for many countries. Finding a method to estimate sodium/salt consumption in national samples leveraging on available data is needed to update and complement the existing evidence.^7–10^ However, quantifying sodium/salt intake based on 24-hour urine samples is costly and burdensome, limiting its use in population-based studies or national health surveys. As an alternative, equations have been developed to estimate sodium/salt intake based on spot urine (SU) samples.^11–14^ However, these equations have been used in few WHO STEPS and other national health surveys,^15^ leaving several countries without data to quantify the local sodium/salt consumption because they do not have access to SU samples.^16^

If we could (accurately) estimate sodium/salt intake based on variables that are routinely available in national health surveys (e.g., weight or blood pressure), mean sodium/salt intake in countries that currently lack urine data (i.e., 24-hour or spot) could be computed using these available predictors. Advanced analytic techniques like machine learning (ML) could make accurate predictions, and inform about the population sodium/salt intake in LMICs. We developed a ML predictive model to estimate salt intake using routinely available variables in WHO STEPS surveys, and we applied this ML model to WHO STEPS surveys without urine data to compute the mean salt intake in the general population.

## METHODS

### Study design

This is a data pooling ML analysis of individual-level data of WHO STEPS surveys. First, we downloaded 19 WHO STEPS surveys with SU samples; these surveys were used for the training, validation and testing the ML model. These 19 WHO STEPS surveys represented 17 LMICs; two countries contributed with two surveys: Bhutan 2014 and 2019 as well as Mongolia 2013 and 2019. Second, we downloaded 49 new WHO STEPS surveys which had the variables included in the ML prediction model (see Variables section), but did not have SU samples. The ML model herein developed was applied to these 49 surveys to estimate the mean salt consumption in the population. All WHO STEPS surveys herein analysed were national (i.e., sub-national surveys were excluded).

### Rationale

We hypothesized that a ML model could accurately predict salt consumption at the individual level, to then inform the overall mean in the underlying population. In addition, we endeavoured to develop a ML model with simple predictors; that is, variables that are routinely available in national health surveys contrary to urine sample that are seldom collected in national health surveys in LMICs. If the model were indeed accurate, then it could be applied to national surveys without urine samples but with the relevant predictors to inform about the mean salt consumption in the target population. These model-driven estimates could be preliminary, until a national health survey is conducted to study mean salt consumption with urine samples (ideally 24-hour urine samples).

### Variables

The predictors we used in the ML model were: sex, age (years), weight (kg), height (m), systolic blood pressure (SBP, mmHg) and diastolic blood pressure (DBP, mmHg).

The WHO STEPS surveys collect anthropometric and three blood pressure measurements. These are taken by trained fieldworkers following a standard protocol.^15^ We used measured weight and height to compute the body mass index (BMI, kg/m^2^). We used the mean SBP and mean DBP of the second and third blood pressure measurements (i.e., the first blood pressure measurement was discarded).

The outcome was salt intake as per the INTERSALT equation.^11^ We chose this equation because it has been used by some WHO STEPS surveys. There is a specific INTERSALT equation for each sex, and they both include the following variables: age (years), BMI (kg/m^2^), SU sodium (mmol/L), and SU creatinine (mmol/L).^11^ We used the following sex-specific formulas.

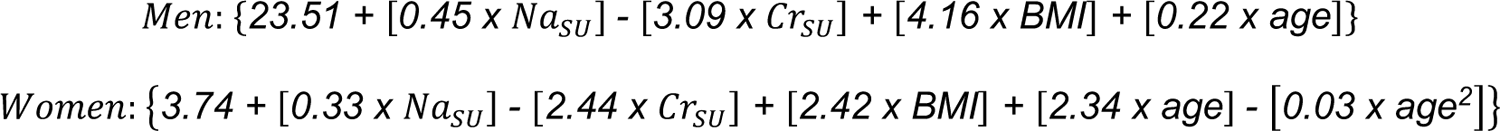

Where the subscript *SU* indicates spot urine, *Na* is sodium, *Cr* is creatinine and *BMI* is body mass index. Because some STEPS surveys had SU creatinine in mg/dl, these values were multiplied by 0.00884 to obtain SU creatinine in mmol/L. No conversion was needed for sodium in SU samples because all WHO STEPS surveys herein included already had urinary sodium in mmol/L. The INTERSALT equation computes 24-hour sodium intake, which is then divided by 17.1 to obtain the salt intake in grams per day (g/d).^11^ For descriptive purposes, we also computed salt intake based on the Kawasaki,^12^ Toft^13^ and Tanaka^14^ equations.

### Analysis

### Data preparation

Our complete-case analysis was restricted to men and non-pregnant women aged between 15 and 69 years. We dropped participants with implausible BMI levels (outside the range 10-80 kg/m^2^) or with implausible weight (outside the range 12-300 kg) or height records (outside the range 1.00-2.50 m). Participants with SBP outside the range 70-270 mmHg were discarded, and so were participants with DBP outside the range 30-150 mmHg. We excluded records with SU creatinine <1.8 or >32.7 mmol/L for males and <1.8 or >28.3 for females.^17, 18^ In addition, we excluded participants with estimated salt intake (using the 4 equations) above or below three standard deviations from the equation-specific mean (Supplementary Figure 1).^19^ After completing data preparation, observations were randomly assigned from the pooled dataset (100%) into three datasets for the ML analysis: training data (50%), test data (30%) and validation data (20%).

### Machine learning modelling

Our research aim was a regression problem where we had a known outcome attribute (salt consumption at the subject level). Therefore, we planned a supervised ML regression analysis. Details about the modelling process are available in the Extended Methods (Supplementary Material pp. 03-06). In brief, we designed a work pipeline with five steps. First, *data analysis*, where we dropped missing observations, we explored the available data to choose scaling and transformation methods to secure all variables were in the same scale or units, and we also planned transformations for categorical variables (e.g., one-hot encoding). Second, *feature importance analysis*, where we investigated the contribution of each predictor to the regression model through methods like Random Forest and Recursive Feature Elimination. The aim of this second step was to exclude any predictor that would not contribute to the regression model. Notably, all predictors (see Variables section) chosen following expert knowledge were kept in the analysis (i.e., the feature importance analysis did not suggest the exclusion of any predictor). Third, *data processing*, having explored the available data (first step in the work pipeline), we implemented different scaling and transformation methods (e.g., Box-Cox, Principal Component Analysis and polynomial features). Fourth, *data modelling*, where we implemented ten ML algorithms: i) Linear Regression (LiR); ii) Hubber Regressor (HuR); iii) Ridge Regressor (RiR); iv) Multilayer Perceptron (MLP); v) Support Vector Regressor (SVR); vi) *k*-Nearest Neighbors (KNN); vii) Random Forest (RF); viii) Gradient Boost Machine (GBM); ix) Extreme Gradient Boosting (XBG); and x) a customized neural network. All these ML algorithms performed similarly, so the decision to choose one was postponed to the fifth (last) step in the work pipeline. Up to this point, we used the training and validations datasets. Five, *forecasting* of the predicted attribute in new data (i.e., data not used for model training); in this step we used the test dataset to choose the model that yielded predictions closest to the observed salt intake. Results comparing the observed and the predicted salt intake were computed in the test dataset alone. For each country we ran a paired t-test between the observed and predicted salt consumption, where a difference was deemed significant at a p <0.05. We also computed the absolute difference between the observed and predicted salt intake. We chose the RF algorithm because it showed the mean difference closest to zero in both sexes combined (observed – predicted = 0) (Supplementary table 1, Supplementary figure 2). All summary estimates (e.g., mean salt intake) were computed accounting for the complex survey design of the WHO STEPS surveys.

### Application of the developed ML model

Having developed the ML model following the steps above described, we applied the model to 49 WHO STEPS national surveys which did not have urine samples but included the predictors in the ML model (see Variables section). In each of these 49 surveys we computed the mean daily salt intake accounting for the complex survey design. These surveys were pre-processed following the same procedures described in the Data preparation section.

### Ethics

We did not seek approval by an Institutional Review Board. We used individual-level survey data which do not include any personal identifiers.

### Role of the funding source

The funder had no role in the study design, analysis, interpretation or decision to publish. The authors are collectively responsible for the accuracy of the data. The arguments and opinions in this work are those of the authors alone, and do not represent the position of the institutions to which they belong.

## RESULTS

### Study population for model derivation and validation

The pooled dataset included 45,152 people from 17 LMICs in 19 WHO STEPS surveys (i.e., two countries, Bhutan and Mongolia, had 2 surveys) conducted between 2013-2019 (Supplementary table 2). Overall, the mean age was 38.6 (95% confidence interval (95% CI): 38.1-39.0) years and the proportion of women was 52.1%. The mean SBP was 122.0 mmHg (95% CI: 121.4-122.6 mmHg) and mean DBP was 79.2 mmHg (95% CI: 78.8-79.6 mmHg). The mean weight was 59.2 kg (95% CI: 58.8-59.7 kg) and the mean height was 1.59 m (95% CI: 1.58-1.59 m).

### Observed and predicted mean salt intake during the ML model derivation and validation

In the test dataset including 19 WHO STEPS surveys, the observed mean salt intake computed as per the INTERSALT equation was 8.1 g/day (95% CI: 8.0-8.2 g/day) across countries. The observed salt intake was higher in men (8.9 g/day; 95% CI: 8.7-9.0 g/day) than in women (7.4 g/day; 95% CI: 7.3-7.4 g/day). Across countries, the predicted mean salt intake was 8.1 g/day (95% CI: 8.1-8.2 g/day). Men had a higher predicted mean salt intake (8.9 g/day; 95% CI: 8.8-8.9 g/day) than women (7.4 g/day; 95% CI: 7.4-7.5 g/day). Overall, there were no substantial differences between the observed and predicted estimates (p=0.065). Results for each survey are presented in Figure 1 and Supplementary table 3.

**Figure 1.**
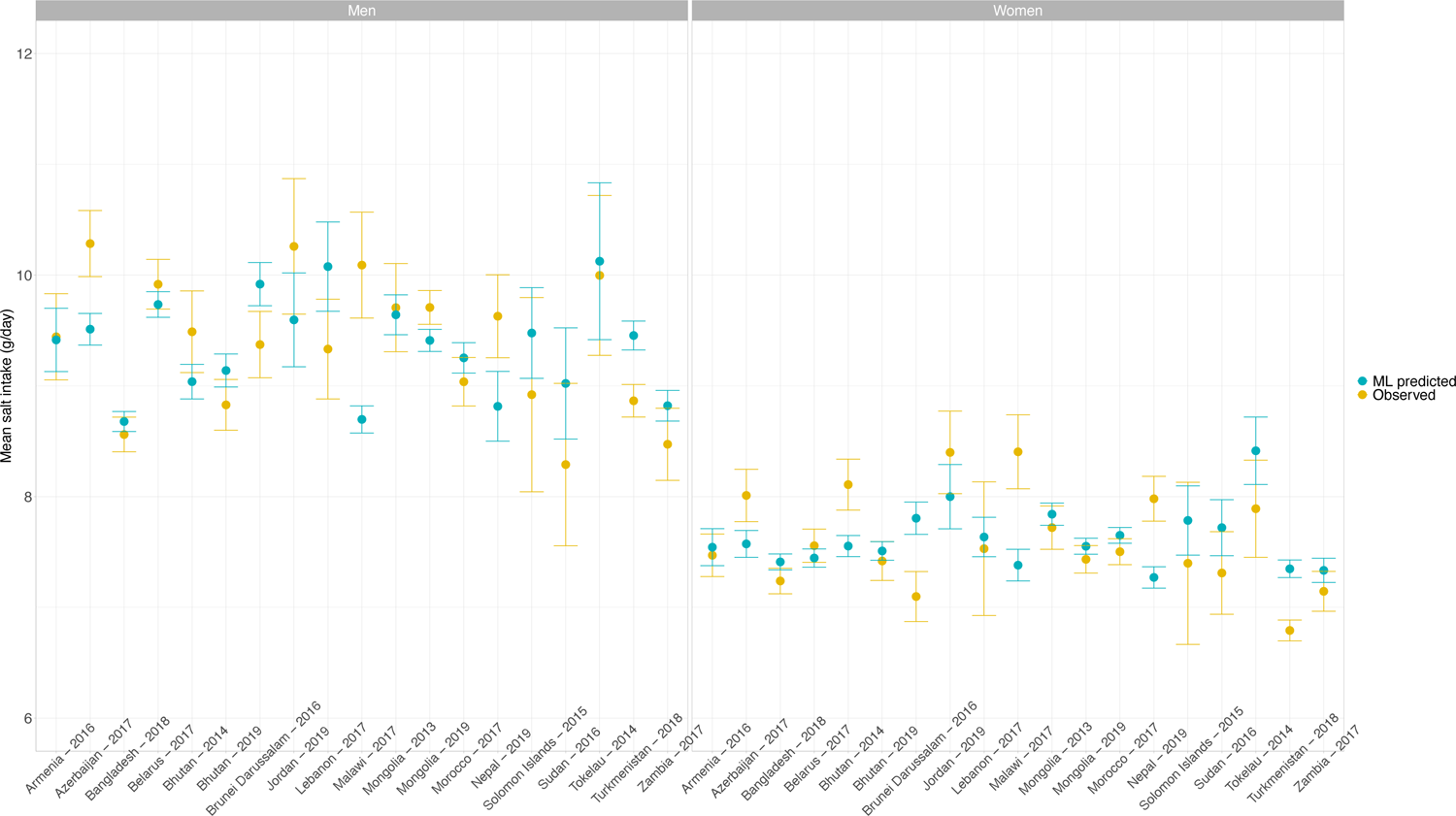
Observed and predicted mean salt intake (g/day) by sex in each survey included in the ML model development Exact estimates (along with their 95% CI) are presented in Supplementary table 3. These results were computed with the test dataset only. Results are for the Random Forest algorithm, which was the model with the best performance.

In men across all countries in the test dataset including 19 WHO STEPS surveys representing 17 LMICs, the mean difference between observed and predicted mean salt intake was 0.02 g/day (p=0.277). Across all surveys, the positive mean difference farthest from zero was 1.39 g/day (Malawi, p<0.001), and the negative mean difference farthest from zero was −0.74 g/day (Lebanon, p=0.227). The mean difference closest to zero was 0.03 g/day (Armenia, p=0.787) (Supplementary table 4).

In women across all countries in the test dataset including 19 WHO STEPS surveys representing 17 LMICs, the mean difference between the observed and predicted mean salt intake was −0.02 g/day (p=0.124). The positive mean difference farthest from zero was 1.02 g/day (Malawi, p<0.001) and the negative mean difference farthest from zero was in −0.71 g/day (Brunei Darussalam, p<0.001). The mean difference closest to zero was −0.07 g/day (Armenia, p=0.343) (Supplementary table 4).

None of the LMICs herein analysed, regardless of the method of sodium intake assessment (i.e., observed or predicted), showed a mean salt intake below the WHO recommended level of <5 g/day (Figure 1, Supplementary table 3). The same occurred for the mean salt intake estimates using the Kawasaki, Toft and Tanaka formulas (Supplementary table 5).

### Implementation of the developed ML model to predict salt consumption in 49 LMICs

The pooled dataset where we applied the ML model included 157,699 people from 49 LMICs in 49 WHO STEPS surveys conducted between 2004-2018 (Supplementary table 6). Overall, the mean age was 37.7 (95% CI: 37.2-38.1) years and the proportion of women was 48.8%. The mean SBP was 123.8 mmHg (95% CI: 123.1-124.4 mmHg) and mean DBP was 79.1 mmHg (95% CI: 78.7-79.6 mmHg). The mean weight was 60.6 kg (95% CI: 60.2-61.0 kg) and the mean height was 1.61 m (95% CI: 1.61-1.61 m).

Across the 49 LMICs, the predicted mean salt intake was 8.1 g/day (95% CI: 8.1-8.2 g/day), and it was higher in men (8.9 g/day; 95% CI: 8.8-8.9 g/day) than in women (7.4 g/day; 95% CI: 7.3-7.4 g/day). None of the LMICs herein analysed, regardless of sex, showed a predicted mean salt intake below the WHO recommended level of <5 g/day (Figure 2, Supplementary table 7).

**Figure 2.**
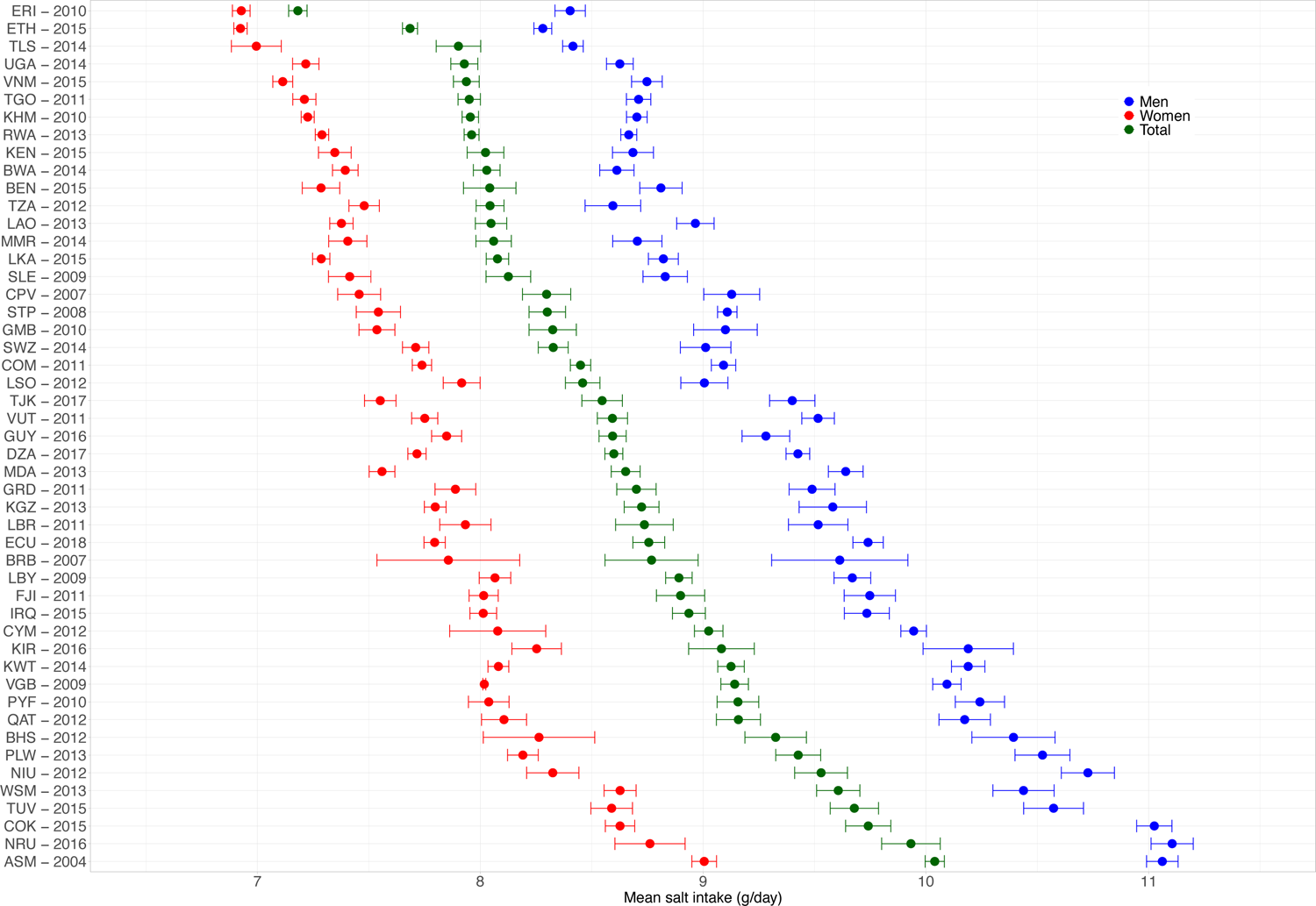
Predicted mean salt intake (g/day) by sex in each of the 49 national surveys included in the application of the model herein developed. Exact estimates (along with their 95% CI) are presented in Supplementary table 7. Countries are presented in ascending order based on their overall mean salt intake (i.e., countries with the highest mean salt intake are at the bottom).

In men, the countries with the highest predicted mean salt intakes were Nauru and American Samoa (both with 11.1 g/day), Cook Islands (11.0 g/day) and Niue (10.7 g/day); remarkably, three of these countries (Nauru, Cook Islands and Niue) are in Western Pacific. In contrast, the lowest predicted mean salt intake in men were in Ethiopia (8.3 g/day), Eritrea and Timor-Leste (both with 8.4 g/day), and United Republic of Tanzania, Botswana and Uganda (all with 8.6 g/day); remarkably, all of these countries except Timor-Leste are in Africa.

In women, the countries with the highest predicted mean salt intakes were American Samoa (9.0 g/day), Nauru (8.8 g/day), and Cook Islands, Samoa and Tuvalu (all with 8.6 g/day); the latter four countries are in Western Pacific. Conversely, the lowest predicted mean salt intake in women were in Ethiopia and Eritrea (both with 6.9 g/day), Timor-Leste (7.0 g/day), and Vietnam (7.1 g/day); the first two countries (out of four) are in Africa.

## DISCUSSION

### Main findings

This work leveraged on 19 national health surveys and readily available predictors to develop a ML model to predict salt consumption; this model was then applied to national surveys in 49 LMICs. The RF ML algorithm yielded the predictions closest to the observed salt intake: the mean difference between predicted and observed salt consumption across surveys was 0.02 g/day in men and −0.02 g/day in women. We used this novel ML model to predict salt consumption in 49 LMICs, where the mean salt consumption ranged from 8.3 g/day (Ethiopia) to 11.1 g/day (Nauru) in men; these numbers in women ranged from 6.9 g/day (Ethiopia and Eritrea) to 9.0 g/day (American Samoa). This work aimed to elaborate on novel analytical tools to predict salt consumption where national surveys have not collected this information limiting their ability to keep track of mean sodium consumption in the general population. Pending external independent validation, our model could be used in monitoring frameworks of salt consumption because most countries do not collect sodium samples in their national health surveys. Our model could contribute to the global surveillance of salt consumption, a relevant cardio-metabolic risk factor.^1–3^

### Public health implications

ML models have been used extensively to predict relevant clinical outcomes (e.g., mortality) and epidemiological indicators (e.g., forecasting COVID-19 cases).^20–24^ Furthermore, ML algorithms have proven to be useful for understanding complex outcomes (e.g., identifying clusters of people with diabetes) based on simple predictors (e.g., BMI) in nationally-representative survey data.^25–27^ Our work complements the current evidence on ML algorithms by demonstrating its use in a relevant field: population salt consumption. In so doing, we delivered a pragmatic tool which could be used to inform the surveillance of salt consumption in countries where national surveys do not objectively collect this information (e.g., SU samples). Moreover, this work provided preliminary evidence to update the global estimates of population-based sodium consumption,^7^ by informing about the mean sodium consumption in 49 LMICs. Our results suggest that mean salt consumption is above the WHO recommended level in all the 49 LMICs herein analysed, and it was the highest among LMICs in Western Pacific, and the lowest among LMICs in Africa. This finding, which is consistent with a global work,^7^ calls for urgent actions to reduce salt consumption in these 49 LMICs, especially those in Western Pacific.

We do not believe that our -or any other-ML model should replace a comprehensive population-based nationally representative health survey with 24-hour or spot urine samples. However, until such surveys are available in many LMICs and periodically conducted, we could suggest using an estimation approach to shed lights about the mean salt consumption in the population. Our ML model seems to be a reasonably good alternative, and could become a pragmatic tool for surveillance systems that keep track of sodium consumption in accordance with global goals.^2, 6^

### Research in context

A global effort provided mean sodium/salt consumption estimates for 187 countries in 1990 and 2010;^7^ they used 24-hour urine samples and dietary reports from surveys conducted in 66 countries, including LMICs. Unfortunately, their results were until 2010 and many LMICs were not included. Our results advanced this evidence by providing more recent salt consumption estimates, because most of the surveys in which we applied our ML model were conducted after 2010 (Supplementary table 6).

Compared to the global estimates for the same LMICs in 2010,^7^ our mean salt consumption estimates were very similar. For example, our 2010 mean salt consumption estimates for Cambodia, Eritrea and the Gambia were 8.0 g/day, 7.2 g/day and 8.3 g/day, whereas the estimates by Powles *et al.* were 11.0 g/day, 5.9 g/day and 7.7 g/day (Supplementary table 8).^7^ We further compared our estimates for surveys conducted between 2007 and 2013 (+/- 3 years around 2010) with the 2010 estimates provided by Powles *et al.*,^7^ and our results were also within reasonable difference. The largest differences were in Kyrgyzstan (8.7 by our ML model versus 13.4 by Powles *et al.*^7^), as well as in Comoros (8.4 versus 4.2^7^) and Rwanda (8.0 versus 4.0^7^). It appears that our predictions were higher than those provided by Powles *et al.*^7^ in countries with presumably low salt consumption (e.g., Comoros, Rwanda); conversely, in countries with presumably high salt consumption (e.g., Kyrgyzstan), our predictions revealed smaller estimates than those by Powles *et al.*^7^ (Supplementary table 8). These differences could be explained by the fact that our ML model was developed based on SU samples rather than 24-hour urine samples as Powles *et al.* did. Strong evidence indicates that estimates based on SU may overestimate salt intake at lower levels of consumption and underestimate salt intake at higher levels of consumption.^28^

In addition to the global work by Powles *et al.*,^7^ there are other reports from some specific LMICs. For example, a survey conducted between 2012 and 2016 with 24-hour urine samples in Fiji and Samoa showed that the mean salt consumption was 10.6 g/day and 7.1 g/day, respectively.^17^ The estimates from our ML model for Fiji (2011) and Samoa (2013) suggested that the mean salt consumption was 8.9 g/day and 9.6 g/day, respectively. A survey in Vanuatu in 2016 based on 24-hour urine sample informed that the mean salt intake was 5.9 g/day;^18^ our estimate for the year 2011was 8.6 g/day. In 2009 in Vietnam a survey with SU samples revealed that the mean salt consumption was 9.9 g/day;^19^ our prediction for the year 2015 was 7.9. These comparisons suggest that our ML-predicted estimates are plausible and close to the best available evidence.

Although these comparisons do not validate our predictions in the 49 national surveys, they suggest that our salt consumption estimates are within reasonable distance from the best available evidence. Until better data are available (e.g., national survey with spot or 24-hour urine sample), our model could provide preliminary evidence to inform the national mean salt consumption. Careful interpretation is warranted to understand the strengths and limitations of our ML-based predictions.

### Strengths and limitations

We followed sound and transparent methods to develop a ML model to predict salt consumption at the individual level. We leveraged on open-access national data collected following standard and consistent protocols (WHO STEPS surveys). Most of the surveys we analysed were conducted after 2010, providing more recent evidence than the latest global effort to quantify salt consumption in all countries.^7^ Notwithstanding, we must acknowledge some limitations. First, urine data was based on a spot sample, which is not the gold standard (24-hour urine sample) to measure daily salt consumption. Future work should verify and advance our results using on 24-hour urine samples available in nationally representative samples; in the meantime, our work has led the foundations and hopefully sparked interest to use available data and novel analytical techniques to deliver estimates of salt consumption in the general population. Second, even though we analysed 19 national surveys (representing 17 LMICs) to develop our ML model, the sample size could still be limited for a data-driven ML algorithm (i.e., 22,577 observations were included in model development). A larger and global work in which all relevant data sources are pooled is needed; while this endeavour takes place, our work has provided recent estimates of salt consumption at the population level in 49 LMICs. In this line, there are still countries which were not herein included. Researchers in these countries, along with local (e.g., ministries of health) and international health authorities (e.g., WHO), should conduct studies/surveys to collect data on salt consumption. This would inform global targets but also local needs and interventions.

### Conclusions

A ML model based on readily available variables was accurate to predict daily salt consumption. This ML model applied to 49 national surveys with no urine samples to compute daily salt consumption, revealed high levels of salt intake particularly in the Western Pacific region. Pending further validation, this ML model could be used to keep track of the overall sodium consumption where resources are not available to conduct national surveys with urine samples.

### DATA SHARING STATEMENT

This study used nationally-representative survey data that are in the public domain, which was requested through the online repository (https://extranet.who.int/ncdsmicrodata/index.php/home). We provide the analysis code of data preparation and data analysis as supplementary materials to this paper.

## SUPPLEMENTARY MATERIAL

### Expanded methods Overview

We worked with a structured dataset which mostly had numeric attributes (variables). Given our study problem, we opted for a supervised learning model because there was a target attribute (i.e., salt consumption at the subject level); specifically, we conducted a supervised regression because the target attribute was a numeric variable. For the machine learning analyses we used Python and the Scikit-Learn library.

First, we developed a pipeline for data management and model development. This way, we followed a consistent and transparent methodology to secure an optimal model for the training set and that would adequately generalize to other (unseen) datasets. The following figure depicts the pipeline we developed: i) we studied the available data and where needed, we did a one-hot encoding; ii) we did feature importance analysis; iii) we chose and tried different scaling and transformation methods, so that all variables would be in the same scale or units; iv) we tried a set of machine learning models, including a customized neural network; v) we forecasted (predicted) the attribute of interest (salt consumption at the subject level) in an unseen dataset (i.e., not used for model training). Notably, we went backwards and forwards (see arrows in the figure) between the four first stages until we reached the best combinations and results for each model. In the following sections, we will describe each of these five stages.

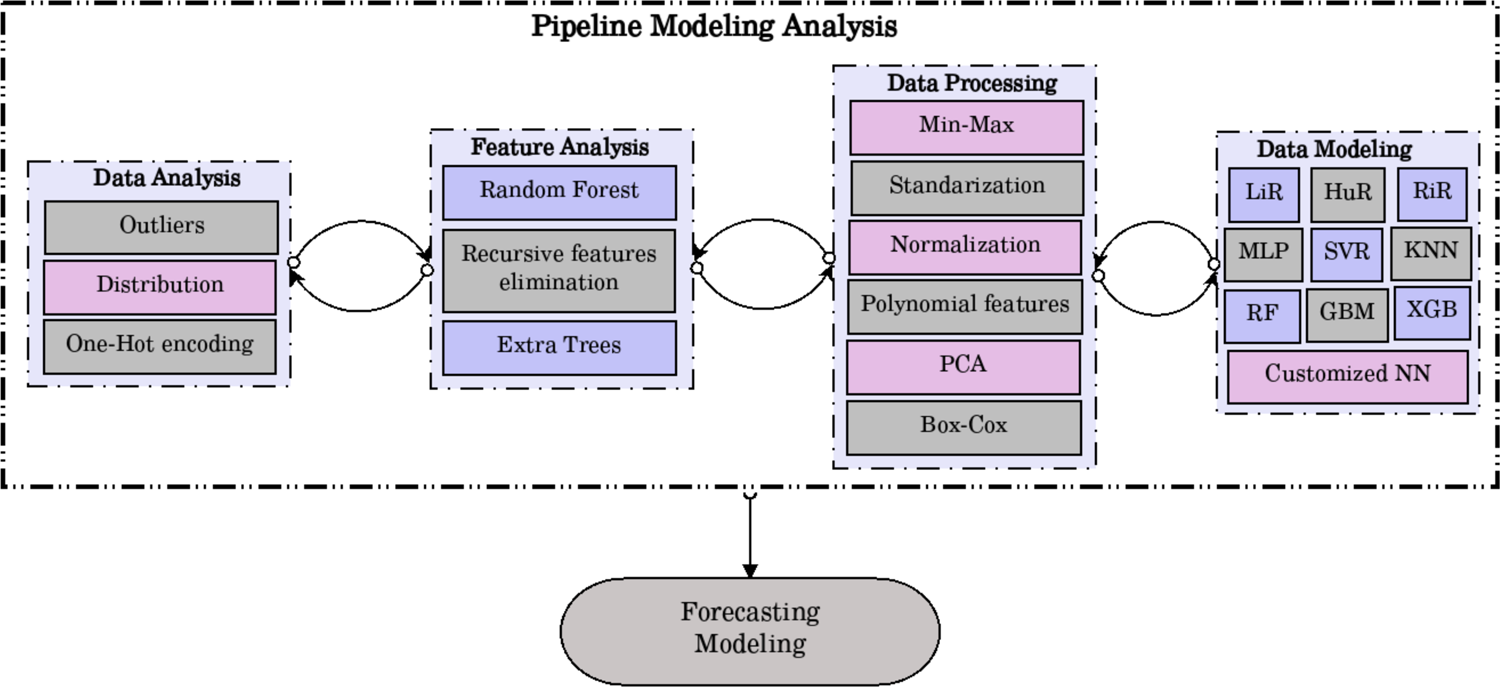

### Data analysis

This was an exploratory analysis to understand the dataset and its characteristics. We worked with a complete-case dataset; in other words, we excluded missing observations in the variables considered in the analysis. Consequently, we did not do any data imputation analysis.

We explored the distribution of all numerical variables, which were in different units and scales; this exploratory analysis informed the choices of data processing methods (e.g., Box-Cox) implemented in the third stage.

### Feature importance analysis

Even though we followed expert knowledge to select a reduced, though relevant number of predictors to be included in the regression model, we conducted feature importance analyses to understand the role each predictor would play in the model. This process aimed to eliminate variables that would not carry substantial information for the model. We used Random Forest, Recursive Feature Elimination and Extra Trees. Consistently, these three methods suggested that all the chosen predictors would contribute to a better model.

### Data processing

As described in the data analysis section (first stage), numeric variables were in different units and scales; therefore, these variables needed to be scaled or transformed. This scaling would also help to find a better prediction model. It is common knowledge that machine learning models would perform differently (and better) depending on data transformation methods. We did: i) Min-Max whereby numeric variables were scaled to a range between 0 and 1; ii) Standardization; iii) Normalization: iv) Polynomial features of degree 2 (quadratic polynomial); v) Principal Component Analysis with 3 components and explained variance of ≥0.95; and vi) Box-Cox.

### Data modelling

There are several machine learning algorithms for a supervised regression model. Those that we used, and that are depicted in the figure, yielded much better results and were studied in detail. That is, at the beginning of our work we explored other algorithms, though these did not perform well and were not considered thereafter. The algorithms we considered were: i) Linear Regression (LiR); ii) Hubber Regressor (HuR); iii) Ridge Regressor (RiR); iv) Multilayer Perceptron (MLP); v) Support Vector Regressor (SVR); vi) *k*-Nearest Neighbors (KNN); vii) Random Forest (RF); viii) Gradient Boost Machine (GBM); and ix) Extreme Gradient Boosting (XBG).

In addition to these nine machine learning algorithms, we also implemented a neural network (see figure below). This neural network was optimized empirically. We used a batch size = 256; epochs = 300; and optimizer = ‘adam’. The neural network was implemented in Python using the Keras library.

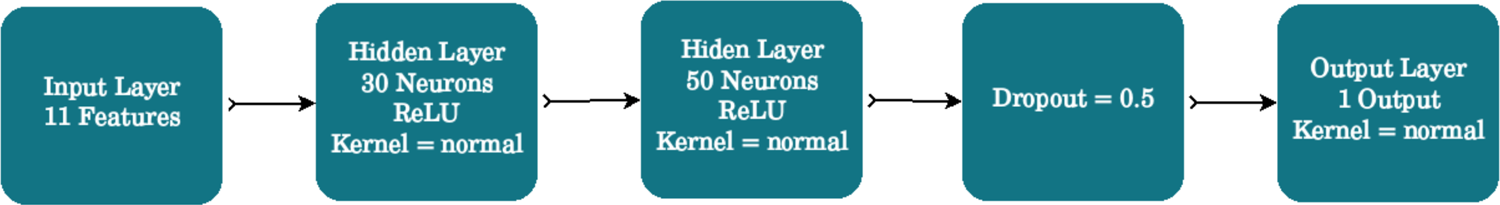

For each model and processing method (see Data Processing section) we studied the R^2^, Mean Absolute Error (MAE) and Root Mean Square Error (RMSE). As shown in the table, all algorithms showed a similar performance. Because all the algorithms had an equivalent performance, the chosen one needed to be defined at the forecasting stage; that is, the one that would generalize better to new (unseen) data.

| Algorithm | Processing | $R^2$ | MAE | RMSE |
| --- | --- | --- | --- | --- |
| LiR | Polynomial(g=2) | 0.442 | 1.1156 | 1.4456 |
| HuB | Polynomial(g=2) | 0.442 | 1.1156 | 1.4455 |
| RiR | Polynomial(g=2) | 0.441 | 1.1163 | 1.4135 |
| MLP | BoxCox | 0.445 | 1.1096 | 1.4418 |
| SVR | MinMax | 0.440 | 1.1000 | 1.4480 |
| KNN | Standarized | 0.416 | 1.1437 | 1.4478 |
| RF | Polynomial(g=2) | 0.422 | 1.1373 | 1.4717 |
| GBM | MinMax | 0.443 | 1.1146 | 1.4432 |
| XGB | BoxCox | 0.424 | 1.1134 | 1.4687 |
| Customized NN | BoxCox | 0.456 | 1.1045 | 1.4445 |

### Forecasting modelling

This stage implies studying the predicted results in new (unseen) data (i.e., data not used for model training). For this stage we used the validation and test datasets. We chose the model that yielded predictions closest to the observed results. In this line, we compared the mean difference between the observed and predicted mean salt intake results (i.e., observed – predicted) across all prediction algorithms.

We observed there was no unique algorithm that had the mean difference closest to zero in men and women at the same time (Supplementary table 1). The RF algorithm had the mean difference closest to zero in both sexes combined (mean difference = 0.0004), the MLP algorithm performed the best in men (mean difference = 0.0052), and in women the GBR algorithm showed the best results (mean difference = −0.0005).

To support our decision process, we plotted the mean differences in men and women for each survey (Supplementary figure 2); this figure only included the predictions based on the top five algorithms (RF, MLP, GBR, LiR and RiR). We counted how many times (i.e., number of surveys) each algorithm had the mean difference closest to zero.

Because the RF algorithm had the mean difference closest to zero in both sexes combined and it was among the top 5 algorithms in men and women (Supplementary table 1), we decided to choose the RF algorithm. Additionally, predictions based on the RF algorithm were the closest to zero across surveys (Supplementary figure 2). These analyses were performed in R (version 4.0.3).

### Algorithm application

To make the predictions in the new 49 datasets without information about urine samples, we used the RF model (i.e., ML algorithm and predictors) developed following the methods above described (see Forecasting Modelling section). We re-trained the model with the full dataset used for model development and validation (i.e., train, validated and test dataset pooled), and then predicted the outcome (i.e., mean salt intake) in the 49 new datasets.

---

**Supplementary figure 1.**
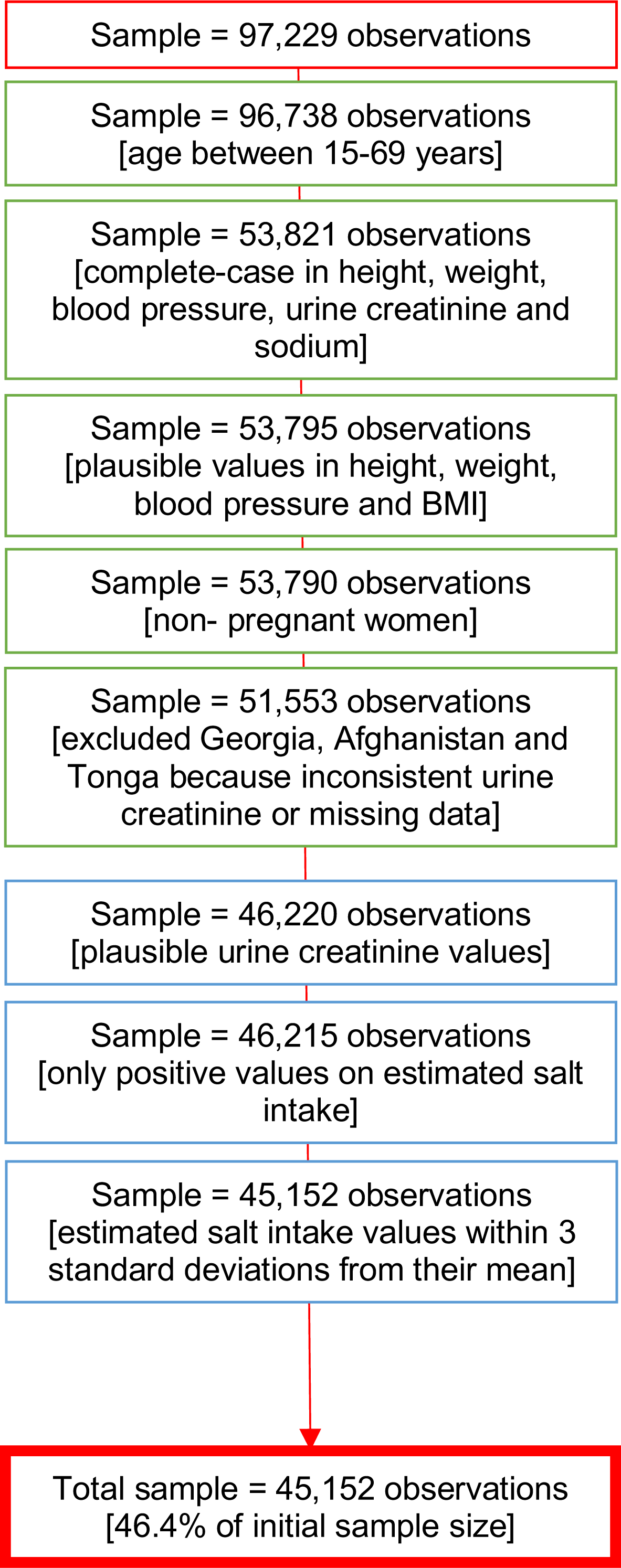
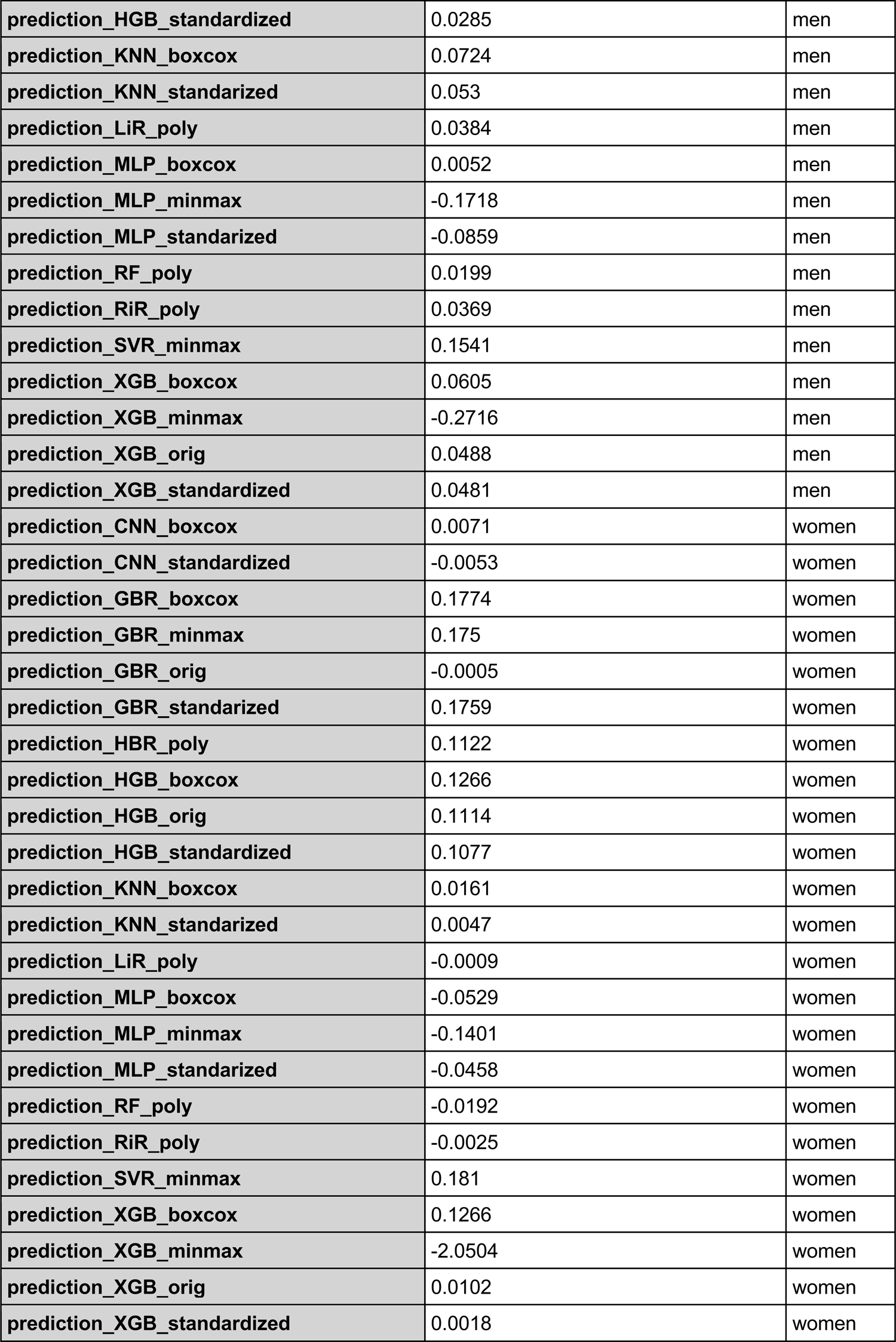
Flowchart of data cleaning and inclusion criteria

**Supplementary Figure 2.**
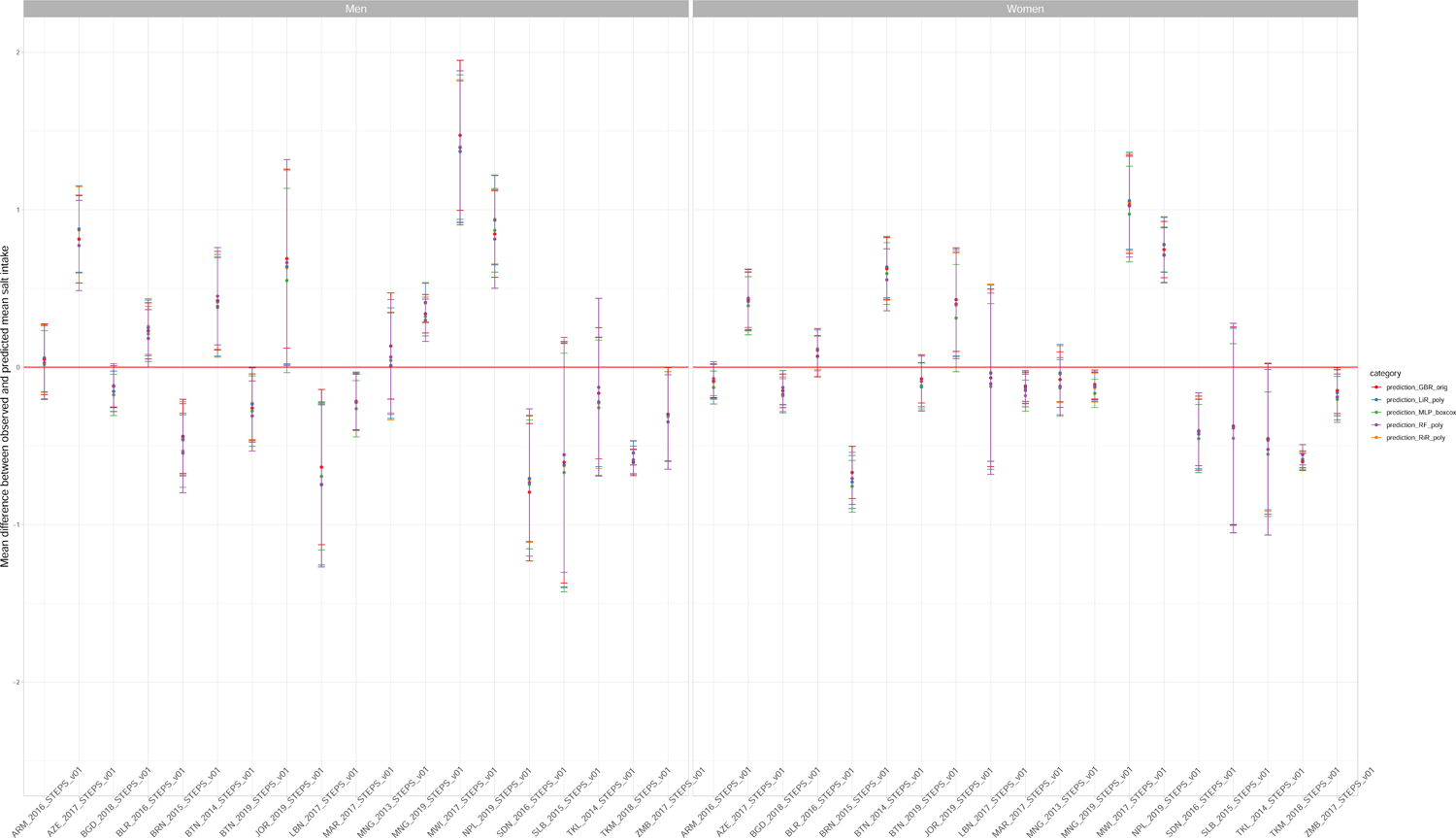
Comparison between mean difference between observed and predicted salt intake across the best algorithms

**Supplementary Table 1.** Mean difference between observed and predicted salt intake by sex across all ML algorithms

| ML algorithm | Mean difference between observed and predicted mean salt intake | Sex |
| --- | --- | --- |
| prediction_CNN_boxcox | 0.0308 | both sexes |
| prediction_CNN_standardized | 0.023 | both sexes |
| prediction_GBR_boxcox | 0.1781 | both sexes |
| prediction_GBR_minmax | 0.1739 | both sexes |
| prediction_GBR_orig | 0.0221 | both sexes |
| prediction_GBR_standarized | 0.1809 | both sexes |
| prediction_HBR_poly | 0.1372 | both sexes |
| prediction_HGB_boxcox | 0.0935 | both sexes |
| prediction_HGB_orig | 0.0676 | both sexes |
| prediction_HGB_standardized | 0.0681 | both sexes |
| prediction_KNN_boxcox | 0.0443 | both sexes |
| prediction_KNN_standarized | 0.0289 | both sexes |
| prediction_LiR_poly | 0.0187 | both sexes |
| prediction_MLP_boxcox | -0.0238 | both sexes |
| prediction_MLP_minmax | -0.1559 | both sexes |
| prediction_MLP_standarized | -0.0659 | both sexes |
| prediction_RF_poly | 0.0004 | both sexes |
| prediction_RiR_poly | 0.0172 | both sexes |
| prediction_SVR_minmax | 0.1675 | both sexes |
| prediction_XGB_boxcox | 0.0935 | both sexes |
| prediction_XGB_minmax | -1.161 | both sexes |
| prediction_XGB_orig | 0.0295 | both sexes |
| prediction_XGB_standardized | 0.0249 | both sexes |
| prediction_CNN_boxcox | 0.0546 | men |
| prediction_CNN_standardized | 0.0514 | men |
| prediction_GBR_boxcox | 0.1788 | men |
| prediction_GBR_minmax | 0.1728 | men |
| prediction_GBR_orig | 0.0447 | men |
| prediction_GBR_standarized | 0.1859 | men |
| prediction_HBR_poly | 0.1621 | men |
| prediction_HGB_boxcox | 0.0605 | men |
| prediction_HGB_orig | 0.0237 | men |
| prediction_HGB_standardized | 0.0285 | men |
| prediction_KNN_boxcox | 0.0724 | men |
| prediction_KNN_standardized | 0.053 | men |
| prediction_LiR_poly | 0.0384 | men |
| prediction_MLP_boxcox | 0.0052 | men |
| prediction_MLP_minmax | -0.1718 | men |
| prediction_MLP_standardized | -0.0859 | men |
| prediction_RF_poly | 0.0199 | men |
| prediction_RiR_poly | 0.0369 | men |
| prediction_SVR_minmax | 0.1541 | men |
| prediction_XGB_boxcox | 0.0605 | men |
| prediction_XGB_minmax | -0.2716 | men |
| prediction_XGB_orig | 0.0488 | men |
| prediction_XGB_standardized | 0.0481 | men |
| prediction_CNN_boxcox | 0.0071 | women |
| prediction_CNN_standardized | -0.0053 | women |
| prediction_GBR_boxcox | 0.1774 | women |
| prediction_GBR_minmax | 0.175 | women |
| prediction_GBR_orig | -0.0005 | women |
| prediction_GBR_standardized | 0.1759 | women |
| prediction_HBR_poly | 0.1122 | women |
| prediction_HGB_boxcox | 0.1266 | women |
| prediction_HGB_orig | 0.1114 | women |
| prediction_HGB_standardized | 0.1077 | women |
| prediction_KNN_boxcox | 0.0161 | women |
| prediction_KNN_standardized | 0.0047 | women |
| prediction_LiR_poly | -0.0009 | women |
| prediction_MLP_boxcox | -0.0529 | women |
| prediction_MLP_minmax | -0.1401 | women |
| prediction_MLP_standardized | -0.0458 | women |
| prediction_RF_poly | -0.0192 | women |
| prediction_RiR_poly | -0.0025 | women |
| prediction_SVR_minmax | 0.181 | women |
| prediction_XGB_boxcox | 0.1266 | women |
| prediction_XGB_minmax | -2.0504 | women |
| prediction_XGB_orig | 0.0102 | women |
| prediction_XGB_standardized | 0.0018 | women |

**Supplementary Table 2.** Weighted distribution of predictors in each survey included in the ML model development.

| Country | Year | Region | Sample size (n) | Mean age (years) | Age range (years) | Proportion of men (%) | Mean, minimum and maximum values of SBP (mmHg) | Mean, minimum and maximum values of DBP (mmHg) | Mean, minimum and maximum values of weight (kg) | Mean, minimum and maximum values of height (m) | Mean, minimum and maximum values of urinary sodium (mmol/L) | Mean, minimum and maximum values of urinary creatinine (mmol/L) |
| --- | --- | --- | --- | --- | --- | --- | --- | --- | --- | --- | --- | --- |
| Armenia | 2016 | Europe | 1074 | 40 | 18-69 | 49.7 | 129.5 (86-237.5) | 84.5 (49-147.5) | 70.9 (35-139) | 1.66 (1.27-1.89) | 128.6 (10.6-237.6) | 10.1 (1.9-27.3) |
| Azerbaijan | 2017 | Europe | 2359 | 39 | 18-69 | 49.5 | 125.9 (81.5-230) | 81.4 (47.5-142.5) | 73.1 (36-174) | 1.67 (1.15-1.98) | 167.7 (2-389) | 11.9 (1.8-31.8) |
| Bangladesh | 2018 | Southeast Asia | 5815 | 39 | 18-69 | 47.4 | 120.2 (72-251) | 78.6 (32-147) | 55.6 (28-111) | 1.56 (1-2.11) | 119.9 (4-403) | 8.4 (2.2-32.3) |
| Belarus | 2017 | Europe | 4506 | 43 | 18-69 | 47.1 | 134.6 (87.5-257) | 84.9 (53.5-147) | 77.7 (41-144) | 1.7 (1.05-1.99) | 149.6 (10.5-371.4) | 12.2 (1.8-32.7) |
| **Bhutan** | 2014 | Southeast Asia | 5734 | 38 | 18-69 | 59.1 | 125.6 (75-228.5) | 84.5 (46-141.5) | 61.4 (23-115) | 1.6 (1.11-1.96) | 142.2 (6-388) | 8.1 (1.9-29.7) |
| **Bhutan** | 2019 | Southeast Asia | 5734 | 34 | 15-69 | 56.5 | 122.9 (85-224.5) | 80.7 (43.5-137) | 61.4 (28.5-140) | 1.58 (1.07-1.92) | 130.5 (4.7-444.9) | 10.4 (1.8-32.7) |
| **Brunei Darussalam** | 2016 | Western Pacific | 1636 | 35 | 18-69 | 51.5 | 122.6 (75.5-218) | 78.4 (45.5-138) | 69 (31.2-138.3) | 1.59 (1.32-1.84) | 122.6 (19.9-329) | 12.6 (1.8-32.6) |
| **Jordan** | 2019 | Eastern Mediterranean | 1040 | 37 | 18-69 | 50.2 | 117.7 (75-200) | 78.4 (50-119.5) | 76.3 (35.5-159.5) | 1.66 (1.36-1.95) | 165.4 (13-365) | 13.6 (1.8-32.5) |
| **Lebanon** | 2017 | Eastern Mediterranean | 999 | 42 | 17-69 | 48.7 | 128.5 (80-214.5) | 76.9 (35-123) | 78.3 (40-141) | 1.68 (1.2-1.96) | 124.4 (4-385) | 11.5 (1.9-32) |
| **Malawi** | 2017 | Africa | 1603 | 35 | 18-69 | 56.3 | 121.8 (74.5-221.5) | 75.6 (39.5-142.5) | 58.4 (33.6-119) | 1.61 (1.36-1.96) | 186.5 (11-399.9) | 10.7 (1.9-32.4) |
| **Mongolia** | 2013 | Western Pacific | 7508 | 42 | 15-64 | 50.3 | 129.3 (88.5-219.5) | 82.1 (49.5-133.5) | 71 (30.6-138) | 1.62 (1.27-1.92) | 134.1 (13.1-515) | 10.9 (1.8-31.9) |
| **Mongolia** | 2019 | Western Pacific | 7508 | 36 | 15-69 | 50.9 | 120.4 (76.5-253.5) | 77.2 (48-143) | 68.5 (29-159) | 1.64 (1.34-1.98) | 117.1 (2.1-348.9) | 7.5 (1.8-28.3) |
| **Morocco** | 2017 | Eastern Mediterranean | 3438 | 40 | 18-69 | 50.6 | 128.1 (83-228.5) | 78.1 (40-139) | 70.9 (35-168) | 1.66 (1.34-1.95) | 122.3 (26.3-575.2) | 10.3 (1.8-31.4) |
| **Nepal** | 2019 | Southeast Asia | 2442 | 36 | 15-69 | 40.7 | 122.8 (81-212.5) | 80.5 (55-146.5) | 54.4 (26-160) | 1.55 (1.21-2.03) | 142.3 (3-437) | 5.6 (1.8-25.5) |
| **Solomon Islands** | 2015 | Western Pacific | 172 | 38 | 18-69 | 61.4 | 121 (88.5-188.5) | 76.5 (52-104.5) | 67.9 (38.5-122) | 1.61 (1.41-1.8) | 99.3 (7-250) | 9.7 (1.9-28.4) |
| **Sudan** | 2016 | Eastern Mediterranean | 571 | 36 | 18-69 | 55.9 | 128.2 (89-231) | 84.9 (58-131.5) | 72.2 (35.6-174) | 1.67 (1.42-1.92) | 128.5 (5-459) | 14 (1.9-32.4) |
| **Tokelau** | 2014 | Western Pacific | 182 | 35 | 18-63 | 56.2 | 124.8 (76-184.5) | 79 (53-128.5) | 94.7 (58-158.3) | 1.71 (1.16-1.88) | 62.9 (20-265) | 5 (2-7.7) |
| **Turkmenistan** | 2018 | Europe | 3584 | 37 | 18-69 | 52.7 | 127.5 (87.5-268) | 82.9 (54.5-149) | 72.4 (39-142) | 1.68 (1.16-1.98) | 109.2 (10-163) | 11.1 (4.5-18.3) |
| **Zambia** | 2017 | Africa | 2489 | 33 | 18-69 | 50.3 | 124.8 (73-248) | 77 (35.5-147.5) | 60.9 (33.8-150) | 1.62 (1.01-2.07) | 137.1 (10-375) | 12.2 (1.8-32.4) |

**Supplementary Table 3.** Observed and predicted mean salt intake (g/day) by sex in each survey included in the ML model development.

| Country | Year | Sex | Mean salt intake (g/day) | Mean salt intake (g/day) lower | Mean salt intake (g/day) upper | Category |
| --- | --- | --- | --- | --- | --- | --- |
| Armenia | 2016 | Men | 9.41 | 9.13 | 9.7 | ML predicted |
| Armenia | 2016 | Men | 9.44 | 9.05 | 9.83 | Observed |
| Armenia | 2016 | Women | 7.54 | 7.38 | 7.71 | ML predicted |
| Armenia | 2016 | Women | 7.47 | 7.28 | 7.66 | Observed |
| Azerbaijan | 2017 | Men | 9.51 | 9.37 | 9.65 | ML predicted |
| Azerbaijan | 2017 | Men | 10.28 | 9.99 | 10.58 | Observed |
| Azerbaijan | 2017 | Women | 7.57 | 7.45 | 7.69 | ML predicted |
| Azerbaijan | 2017 | Women | 8.01 | 7.77 | 8.25 | Observed |
| Bangladesh | 2018 | Men | 8.68 | 8.59 | 8.77 | ML predicted |
| Bangladesh | 2018 | Men | 8.56 | 8.4 | 8.72 | Observed |
| Bangladesh | 2018 | Women | 7.41 | 7.34 | 7.48 | ML predicted |
| Bangladesh | 2018 | Women | 7.24 | 7.12 | 7.35 | Observed |
| Belarus | 2017 | Men | 9.73 | 9.62 | 9.85 | ML predicted |
| Belarus | 2017 | Men | 9.92 | 9.69 | 10.14 | Observed |
| Belarus | 2017 | Women | 7.45 | 7.36 | 7.53 | ML predicted |
| Belarus | 2017 | Women | 7.56 | 7.41 | 7.71 | Observed |
| Bhutan | 2014 | Men | 9.04 | 8.88 | 9.19 | ML predicted |
| Bhutan | 2014 | Men | 9.49 | 9.12 | 9.86 | Observed |
| Bhutan | 2014 | Women | 7.55 | 7.46 | 7.65 | ML predicted |
| Bhutan | 2014 | Women | 8.11 | 7.88 | 8.34 | Observed |
| Bhutan | 2019 | Men | 9.14 | 8.99 | 9.29 | ML predicted |
| Bhutan | 2019 | Men | 8.83 | 8.6 | 9.06 | Observed |
| Bhutan | 2019 | Women | 7.51 | 7.42 | 7.59 | ML predicted |
| Bhutan | 2019 | Women | 7.42 | 7.24 | 7.59 | Observed |
| Brunei Darussalam | 2016 | Men | 9.92 | 9.72 | 10.11 | ML predicted |
| **Brunei Darussalam** | 2016 | Men | 9.37 | 9.07 | 9.67 | Observed |
| **Brunei Darussalam** | 2016 | Women | 7.8 | 7.66 | 7.95 | ML predicted |
| **Brunei Darussalam** | 2016 | Women | 7.1 | 6.87 | 7.32 | Observed |
| **Jordan** | 2019 | Men | 9.6 | 9.17 | 10.02 | ML predicted |
| **Jordan** | 2019 | Men | 10.26 | 9.65 | 10.87 | Observed |
| **Jordan** | 2019 | Women | 8 | 7.71 | 8.29 | ML predicted |
| **Jordan** | 2019 | Women | 8.4 | 8.03 | 8.77 | Observed |
| **Lebanon** | 2017 | Men | 10.08 | 9.67 | 10.48 | ML predicted |
| **Lebanon** | 2017 | Men | 9.33 | 8.88 | 9.78 | Observed |
| **Lebanon** | 2017 | Women | 7.63 | 7.46 | 7.81 | ML predicted |
| **Lebanon** | 2017 | Women | 7.53 | 6.93 | 8.13 | Observed |
| **Malawi** | 2017 | Men | 8.7 | 8.57 | 8.82 | ML predicted |
| **Malawi** | 2017 | Men | 10.09 | 9.61 | 10.57 | Observed |
| **Malawi** | 2017 | Women | 7.38 | 7.24 | 7.52 | ML predicted |
| **Malawi** | 2017 | Women | 8.4 | 8.07 | 8.74 | Observed |
| **Mongolia** | 2013 | Men | 9.64 | 9.46 | 9.82 | ML predicted |
| **Mongolia** | 2013 | Men | 9.71 | 9.31 | 10.1 | Observed |
| **Mongolia** | 2013 | Women | 7.84 | 7.74 | 7.94 | ML predicted |
| **Mongolia** | 2013 | Women | 7.72 | 7.52 | 7.92 | Observed |
| **Mongolia** | 2019 | Men | 9.41 | 9.31 | 9.51 | ML predicted |
| **Mongolia** | 2019 | Men | 9.71 | 9.55 | 9.86 | Observed |
| **Mongolia** | 2019 | Women | 7.55 | 7.48 | 7.62 | ML predicted |
| **Mongolia** | 2019 | Women | 7.43 | 7.31 | 7.56 | Observed |
| **Morocco** | 2017 | Men | 9.25 | 9.12 | 9.39 | ML predicted |
| **Morocco** | 2017 | Men | 9.04 | 8.82 | 9.26 | Observed |
| **Morocco** | 2017 | Women | 7.65 | 7.58 | 7.72 | ML predicted |
| **Morocco** | 2017 | Women | 7.5 | 7.39 | 7.62 | Observed |
| **Nepal** | 2019 | Men | 8.81 | 8.5 | 9.13 | ML predicted |
| **Nepal** | 2019 | Men | 9.63 | 9.25 | 10 | Observed |
| **Nepal** | 2019 | Women | 7.27 | 7.17 | 7.37 | ML predicted |
| **Nepal** | 2019 | Women | 7.98 | 7.78 | 8.18 | Observed |
| **Solomon Islands** | 2015 | Men | 9.48 | 9.07 | 9.89 | ML predicted |
| **Solomon Islands** | 2015 | Men | 8.92 | 8.04 | 9.8 | Observed |
| **Solomon Islands** | 2015 | Women | 7.78 | 7.47 | 8.1 | ML predicted |
| **Solomon Islands** | 2015 | Women | 7.4 | 6.67 | 8.13 | Observed |
| **Sudan** | 2016 | Men | 9.02 | 8.52 | 9.52 | ML predicted |
| **Sudan** | 2016 | Men | 8.29 | 7.56 | 9.02 | Observed |
| **Sudan** | 2016 | Women | 7.72 | 7.47 | 7.97 | ML predicted |
| **Sudan** | 2016 | Women | 7.31 | 6.94 | 7.68 | Observed |
| **Tokelau** | 2014 | Men | 10.13 | 9.42 | 10.83 | ML predicted |
| **Tokelau** | 2014 | Men | 10 | 9.28 | 10.72 | Observed |
| **Tokelau** | 2014 | Women | 8.41 | 8.11 | 8.72 | ML predicted |
| **Tokelau** | 2014 | Women | 7.89 | 7.45 | 8.33 | Observed |
| **Turkmenistan** | 2018 | Men | 9.45 | 9.32 | 9.58 | ML predicted |
| **Turkmenistan** | 2018 | Men | 8.87 | 8.72 | 9.01 | Observed |
| **Turkmenistan** | 2018 | Women | 7.35 | 7.27 | 7.43 | ML predicted |
| **Turkmenistan** | 2018 | Women | 6.79 | 6.7 | 6.89 | Observed |
| **Zambia** | 2017 | Men | 8.82 | 8.68 | 8.96 | ML predicted |
| **Zambia** | 2017 | Men | 8.47 | 8.15 | 8.8 | Observed |
| **Zambia** | 2017 | Women | 7.33 | 7.22 | 7.44 | ML predicted |
| **Zambia** | 2017 | Women | 7.14 | 6.96 | 7.32 | Observed |

**Supplementary Table 4.** Mean difference between observed and predicted salt intake by sex in each survey included in the ML model development.

| Country | Year | Sex | Mean difference (g/day) | Mean difference (g/day) lower | Mean difference (g/day) upper | Category | P value |
| --- | --- | --- | --- | --- | --- | --- | --- |
| Armenia | 2016 | Men | 0.03 | -0.21 | 0.26 | Observed vs predicted | 0.7873 |
| Armenia | 2016 | Women | -0.07 | -0.18 | 0.03 | Observed vs predicted | 0.3432 |
| Azerbaijan | 2017 | Men | 0.77 | 0.49 | 1.06 | Observed vs predicted | <0.0001 |
| Azerbaijan | 2017 | Women | 0.44 | 0.25 | 0.62 | Observed vs predicted | <0.0001 |
| Bangladesh | 2018 | Men | -0.12 | -0.26 | 0.02 | Observed vs predicted | 0.0033 |
| Bangladesh | 2018 | Women | -0.17 | -0.28 | -0.06 | Observed vs predicted | 0.0002 |
| Belarus | 2017 | Men | 0.18 | 0 | 0.37 | Observed vs predicted | 0.0032 |
| Belarus | 2017 | Women | 0.11 | -0.02 | 0.24 | Observed vs predicted | 0.004 |
| Bhutan | 2014 | Men | 0.45 | 0.14 | 0.76 | Observed vs predicted | <0.0001 |
| Bhutan | 2014 | Women | 0.55 | 0.36 | 0.75 | Observed vs predicted | <0.0001 |
| Bhutan | 2019 | Men | -0.31 | -0.53 | -0.09 | Observed vs predicted | <0.0001 |
| Bhutan | 2019 | Women | -0.09 | -0.25 | 0.07 | Observed vs predicted | 0.108 |
| Brunei Darussalam | 2016 | Men | -0.55 | -0.8 | -0.29 | Observed vs predicted | <0.0001 |
| Brunei Darussalam | 2016 | Women | -0.71 | -0.87 | -0.54 | Observed vs predicted | <0.0001 |
| **Jordan** | 2019 | Men | 0.66 | 0.01 | 1.32 | Observed<br>vs<br>predicted | 0.75 |
| **Jordan** | 2019 | Women | 0.4 | 0.05 | 0.75 | Observed<br>vs<br>predicted | 0.0513 |
| **Lebanon** | 2017 | Men | -0.74 | -1.27 | -0.22 | Observed<br>vs<br>predicted | 0.2274 |
| **Lebanon** | 2017 | Women | -0.11 | -0.68 | 0.47 | Observed<br>vs<br>predicted | 0.4273 |
| **Malawi** | 2017 | Men | 1.39 | 0.9 | 1.88 | Observed<br>vs<br>predicted | <0.0001 |
| **Malawi** | 2017 | Women | 1.02 | 0.7 | 1.35 | Observed<br>vs<br>predicted | <0.0001 |
| **Mongolia** | 2013 | Men | 0.06 | -0.3 | 0.43 | Observed<br>vs<br>predicted | 0.4277 |
| **Mongolia** | 2013 | Women | -0.12 | -0.3 | 0.06 | Observed<br>vs<br>predicted | 0.3048 |
| **Mongolia** | 2019 | Men | 0.3 | 0.16 | 0.43 | Observed<br>vs<br>predicted | <0.0001 |
| **Mongolia** | 2019 | Women | -0.12 | -0.21 | -0.03 | Observed<br>vs<br>predicted | 0.0009 |
| **Morocco** | 2017 | Men | -0.22 | -0.4 | -0.03 | Observed<br>vs<br>predicted | 0.0171 |
| **Morocco** | 2017 | Women | -0.15 | -0.25 | -0.04 | Observed<br>vs<br>predicted | 0.0095 |
| **Nepal** | 2019 | Men | 0.81 | 0.5 | 1.13 | Observed<br>vs<br>predicted | <0.0001 |
| **Nepal** | 2019 | Women | 0.71 | 0.54 | 0.88 | Observed<br>vs<br>predicted | <0.0001 |
| **Solomon**<br>**Islands** | 2015 | Men | -0.56 | -1.3 | 0.19 | Observed<br>vs<br>predicted | 0.0358 |
| **Solomon**<br>**Islands** | 2015 | Women | -0.39 | -1.05 | 0.28 | Observed<br>vs<br>predicted | 0.1386 |
| **Sudan** | 2016 | Men | -0.73 | -1.2 | -0.27 | Observed<br>vs<br>predicted | 0.0018 |
| **Sudan** | 2016 | Women | -0.41 | -0.66 | -0.16 | Observed<br>vs<br>predicted | <0.0001 |
| **Tokelau** | 2014 | Men | -0.13 | -0.69 | 0.44 | Observed<br>vs<br>predicted | 0.9361 |
| **Tokelau** | 2014 | Women | -0.52 | -1.07 | 0.02 | Observed<br>vs<br>predicted | 0.279 |
| **Turkmenistan** | 2018 | Men | -0.59 | -0.68 | -0.5 | Observed<br>vs<br>predicted | <0.0001 |
| **Turkmenistan** | 2018 | Women | -0.56 | -0.62 | -0.49 | Observed<br>vs<br>predicted | <0.0001 |
| **Zambia** | 2017 | Men | -0.35 | -0.65 | -0.05 | Observed<br>vs<br>predicted | 0.0004 |
| **Zambia** | 2017 | Women | -0.19 | -0.34 | -0.04 | Observed<br>vs<br>predicted | 0.0001 |

**Supplementary Table 5.** Observed mean salt intake (g/day) by equation and sex in each survey included in the ML model development.

| Country | Year | Sex | Mean salt intake (g/day) | Mean salt intake (g/day) lower | Mean salt intake (g/day) upper | Category |
| --- | --- | --- | --- | --- | --- | --- |
| Armenia | 2016 | Men | 9.44 | 9.05 | 9.83 | Observed_intersalt |
| Armenia | 2016 | Men | 13.79 | 13.03 | 14.55 | Observed_kawasaki |
| Armenia | 2016 | Men | 9.87 | 9.43 | 10.3 | Observed_tanaka |
| Armenia | 2016 | Men | 12.31 | 11.84 | 12.78 | Observed_toft |
| Armenia | 2016 | Women | 7.47 | 7.28 | 7.66 | Observed_intersalt |
| Armenia | 2016 | Women | 12.18 | 11.67 | 12.69 | Observed_kawasaki |
| Armenia | 2016 | Women | 9.81 | 9.47 | 10.15 | Observed_tanaka |
| Armenia | 2016 | Women | 8.34 | 8.2 | 8.47 | Observed_toft |
| Azerbaijan | 2017 | Men | 10.28 | 9.99 | 10.58 | Observed_intersalt |
| Azerbaijan | 2017 | Men | 14.92 | 14.28 | 15.55 | Observed_kawasaki |
| Azerbaijan | 2017 | Men | 10.37 | 10.02 | 10.72 | Observed_tanaka |
| Azerbaijan | 2017 | Men | 12.88 | 12.5 | 13.25 | Observed_toft |
| Azerbaijan | 2017 | Women | 8.01 | 7.77 | 8.25 | Observed_intersalt |
| Azerbaijan | 2017 | Women | 12.48 | 11.92 | 13.03 | Observed_kawasaki |
| Azerbaijan | 2017 | Women | 10.05 | 9.7 | 10.4 | Observed_tanaka |
| Azerbaijan | 2017 | Women | 8.4 | 8.25 | 8.55 | Observed_toft |
| Bangladesh | 2018 | Men | 8.56 | 8.4 | 8.72 | Observed_intersalt |
| Bangladesh | 2018 | Men | 12.9 | 12.61 | 13.19 | Observed_kawasaki |
| Bangladesh | 2018 | Men | 8.94 | 8.77 | 9.1 | Observed_tanaka |
| Bangladesh | 2018 | Men | 11.81 | 11.63 | 11.99 | Observed_toft |
| Bangladesh | 2018 | Women | 7.24 | 7.12 | 7.35 | Observed_intersalt |
| Bangladesh | 2018 | Women | 11.91 | 11.62 | 12.19 | Observed_kawasaki |
| Bangladesh | 2018 | Women | 8.91 | 8.73 | 9.08 | Observed_tanaka |
| Bangladesh | 2018 | Women | 8.28 | 8.2 | 8.36 | Observed_toft |
| Belarus | 2017 | Men | 9.92 | 9.69 | 10.14 | Observed_intersalt |
| Belarus | 2017 | Men | 13.88 | 13.51 | 14.25 | Observed_kawasaki |
| Belarus | 2017 | Men | 9.97 | 9.76 | 10.19 | Observed_tanaka |
| Belarus | 2017 | Men | 12.24 | 12.01 | 12.47 | Observed_toft |
| Belarus | 2017 | Women | 7.56 | 7.41 | 7.71 | Observed_intersalt |
| Belarus | 2017 | Women | 11.61 | 11.31 | 11.92 | Observed_kawasaki |
| Belarus | 2017 | Women | 9.67 | 9.47 | 9.87 | Observed_tanaka |
| Belarus | 2017 | Women | 8.14 | 8.06 | 8.23 | Observed_toft |
| Bhutan | 2014 | Men | 9.49 | 9.12 | 9.86 | Observed_intersalt |
| **Bhutan** | 2014 | Men | 14.44 | 13.65 | 15.23 | Observed_kawasaki |
| **Bhutan** | 2014 | Men | 9.77 | 9.33 | 10.2 | Observed_tanaka |
| **Bhutan** | 2014 | Men | 12.68 | 12.2 | 13.16 | Observed_toft |
| **Bhutan** | 2014 | Women | 8.11 | 7.88 | 8.34 | Observed_intersalt |
| **Bhutan** | 2014 | Women | 14.11 | 13.57 | 14.66 | Observed_kawasaki |
| **Bhutan** | 2014 | Women | 10.53 | 10.19 | 10.87 | Observed_tanaka |
| **Bhutan** | 2014 | Women | 8.81 | 8.67 | 8.95 | Observed_toft |
| **Bhutan** | 2019 | Men | 8.83 | 8.6 | 9.06 | Observed_intersalt |
| **Bhutan** | 2019 | Men | 12.34 | 11.86 | 12.82 | Observed_kawasaki |
| **Bhutan** | 2019 | Men | 8.51 | 8.27 | 8.75 | Observed_tanaka |
| **Bhutan** | 2019 | Men | 11.36 | 11.06 | 11.65 | Observed_toft |
| **Bhutan** | 2019 | Women | 7.42 | 7.24 | 7.59 | Observed_intersalt |
| **Bhutan** | 2019 | Women | 11.63 | 11.23 | 12.02 | Observed_kawasaki |
| **Bhutan** | 2019 | Women | 8.91 | 8.66 | 9.15 | Observed_tanaka |
| **Bhutan** | 2019 | Women | 8.2 | 8.09 | 8.3 | Observed_toft |
| **Brunei Darussalam** | 2016 | Men | 9.37 | 9.07 | 9.67 | Observed_intersalt |
| **Brunei Darussalam** | 2016 | Men | 11.95 | 11.52 | 12.38 | Observed_kawasaki |
| **Brunei Darussalam** | 2016 | Men | 8.44 | 8.2 | 8.68 | Observed_tanaka |
| **Brunei Darussalam** | 2016 | Men | 11.09 | 10.82 | 11.36 | Observed_toft |
| **Brunei Darussalam** | 2016 | Women | 7.1 | 6.87 | 7.32 | Observed_intersalt |
| **Brunei Darussalam** | 2016 | Women | 10 | 9.62 | 10.38 | Observed_kawasaki |
| **Brunei Darussalam** | 2016 | Women | 8.1 | 7.85 | 8.36 | Observed_tanaka |
| **Brunei Darussalam** | 2016 | Women | 7.75 | 7.63 | 7.86 | Observed_toft |
| **Jordan** | 2019 | Men | 10.26 | 9.65 | 10.87 | Observed_intersalt |
| **Jordan** | 2019 | Men | 13.76 | 12.59 | 14.93 | Observed_kawasaki |
| **Jordan** | 2019 | Men | 9.71 | 9.12 | 10.3 | Observed_tanaka |
| **Jordan** | 2019 | Men | 12.18 | 11.51 | 12.84 | Observed_toft |
| **Jordan** | 2019 | Women | 8.4 | 8.03 | 8.77 | Observed_intersalt |
| **Jordan** | 2019 | Women | 12.28 | 11.58 | 12.98 | Observed_kawasaki |
| **Jordan** | 2019 | Women | 9.81 | 9.36 | 10.26 | Observed_tanaka |
| **Jordan** | 2019 | Women | 8.37 | 8.18 | 8.57 | Observed_toft |
| **Lebanon** | 2017 | Men | 9.33 | 8.88 | 9.78 | Observed_intersalt |
| **Lebanon** | 2017 | Men | 12.79 | 11.35 | 14.23 | Observed_kawasaki |
| Lebanon | 2017 | Men | 9.21 | 8.41 | 10 | Observed_tanaka |
| Lebanon | 2017 | Men | 11.5 | 10.6 | 12.4 | Observed_toft |
| Lebanon | 2017 | Women | 7.53 | 6.93 | 8.13 | Observed_intersalt |
| Lebanon | 2017 | Women | 11.48 | 10.08 | 12.88 | Observed_kawasaki |
| Lebanon | 2017 | Women | 9.44 | 8.44 | 10.44 | Observed_tanaka |
| Lebanon | 2017 | Women | 8.06 | 7.58 | 8.54 | Observed_toft |
| Malawi | 2017 | Men | 10.09 | 9.61 | 10.57 | Observed_intersalt |
| Malawi | 2017 | Men | 14.37 | 13.72 | 15.02 | Observed_kawasaki |
| Malawi | 2017 | Men | 9.62 | 9.26 | 9.98 | Observed_tanaka |
| Malawi | 2017 | Men | 12.68 | 12.3 | 13.07 | Observed_toft |
| Malawi | 2017 | Women | 8.4 | 8.07 | 8.74 | Observed_intersalt |
| Malawi | 2017 | Women | 13.74 | 12.87 | 14.6 | Observed_kawasaki |
| Malawi | 2017 | Women | 10.32 | 9.8 | 10.84 | Observed_tanaka |
| Malawi | 2017 | Women | 8.69 | 8.48 | 8.91 | Observed_toft |
| Mongolia | 2013 | Men | 9.71 | 9.31 | 10.1 | Observed_intersalt |
| Mongolia | 2013 | Men | 13.25 | 12.63 | 13.87 | Observed_kawasaki |
| Mongolia | 2013 | Men | 9.37 | 9.04 | 9.7 | Observed_tanaka |
| Mongolia | 2013 | Men | 11.96 | 11.57 | 12.35 | Observed_toft |
| Mongolia | 2013 | Women | 7.72 | 7.52 | 7.92 | Observed_intersalt |
| Mongolia | 2013 | Women | 11.65 | 11.11 | 12.2 | Observed_kawasaki |
| Mongolia | 2013 | Women | 9.37 | 9.01 | 9.73 | Observed_tanaka |
| Mongolia | 2013 | Women | 8.18 | 8.03 | 8.34 | Observed_toft |
| Mongolia | 2019 | Men | 9.71 | 9.55 | 9.86 | Observed_intersalt |
| Mongolia | 2019 | Men | 14.77 | 14.43 | 15.1 | Observed_kawasaki |
| Mongolia | 2019 | Men | 10.09 | 9.91 | 10.27 | Observed_tanaka |
| Mongolia | 2019 | Men | 12.81 | 12.61 | 13 | Observed_toft |
| Mongolia | 2019 | Women | 7.43 | 7.31 | 7.56 | Observed_intersalt |
| Mongolia | 2019 | Women | 11.97 | 11.68 | 12.25 | Observed_kawasaki |
| Mongolia | 2019 | Women | 9.54 | 9.35 | 9.73 | Observed_tanaka |
| Mongolia | 2019 | Women | 8.27 | 8.19 | 8.35 | Observed_toft |
| Morocco | 2017 | Men | 9.04 | 8.82 | 9.26 | Observed_intersalt |
| Morocco | 2017 | Men | 13.14 | 12.7 | 13.58 | Observed_kawasaki |
| Morocco | 2017 | Men | 9.41 | 9.16 | 9.66 | Observed_tanaka |
| Morocco | 2017 | Men | 11.82 | 11.55 | 12.09 | Observed_toft |
| Morocco | 2017 | Women | 7.5 | 7.39 | 7.62 | Observed_intersalt |
| Morocco | 2017 | Women | 11.74 | 11.43 | 12.04 | Observed_kawasaki |
| Morocco | 2017 | Women | 9.5 | 9.3 | 9.69 | Observed_tanaka |
| Morocco | 2017 | Women | 8.19 | 8.11 | 8.27 | Observed_toft |
| **Nepal** | 2019 | Men | 9.63 | 9.25 | 10 | Observed_intersalt |
| **Nepal** | 2019 | Men | 16.78 | 15.86 | 17.69 | Observed_kawasaki |
| **Nepal** | 2019 | Men | 10.82 | 10.34 | 11.29 | Observed_tanaka |
| **Nepal** | 2019 | Men | 14.15 | 13.63 | 14.68 | Observed_toft |
| **Nepal** | 2019 | Women | 7.98 | 7.78 | 8.18 | Observed_intersalt |
| **Nepal** | 2019 | Women | 15.76 | 15.21 | 16.32 | Observed_kawasaki |
| **Nepal** | 2019 | Women | 11.15 | 10.83 | 11.46 | Observed_tanaka |
| **Nepal** | 2019 | Women | 9.21 | 9.09 | 9.34 | Observed_toft |
| **Solomon Islands** | 2015 | Men | 8.92 | 8.04 | 9.8 | Observed_intersalt |
| **Solomon Islands** | 2015 | Men | 12.07 | 10.21 | 13.92 | Observed_kawasaki |
| **Solomon Islands** | 2015 | Men | 8.63 | 7.55 | 9.7 | Observed_tanaka |
| **Solomon Islands** | 2015 | Men | 11.14 | 9.94 | 12.33 | Observed_toft |
| **Solomon Islands** | 2015 | Women | 7.4 | 6.67 | 8.13 | Observed_intersalt |
| **Solomon Islands** | 2015 | Women | 13.18 | 10.5 | 15.86 | Observed_kawasaki |
| **Solomon Islands** | 2015 | Women | 9.96 | 8.3 | 11.63 | Observed_tanaka |
| **Solomon Islands** | 2015 | Women | 8.44 | 7.74 | 9.15 | Observed_toft |
| **Sudan** | 2016 | Men | 8.29 | 7.56 | 9.02 | Observed_intersalt |
| **Sudan** | 2016 | Men | 11.27 | 10.41 | 12.13 | Observed_kawasaki |
| **Sudan** | 2016 | Men | 8.19 | 7.66 | 8.73 | Observed_tanaka |
| **Sudan** | 2016 | Men | 10.6 | 10.04 | 11.15 | Observed_toft |
| **Sudan** | 2016 | Women | 7.31 | 6.94 | 7.68 | Observed_intersalt |
| **Sudan** | 2016 | Women | 10.36 | 9.8 | 10.93 | Observed_kawasaki |
| **Sudan** | 2016 | Women | 8.72 | 8.31 | 9.12 | Observed_tanaka |
| **Sudan** | 2016 | Women | 7.81 | 7.65 | 7.98 | Observed_toft |
| **Tokelau** | 2014 | Men | 10 | 9.28 | 10.72 | Observed_intersalt |
| **Tokelau** | 2014 | Men | 15.6 | 14.19 | 17 | Observed_kawasaki |
| **Tokelau** | 2014 | Men | 10.89 | 10.05 | 11.73 | Observed_tanaka |
| **Tokelau** | 2014 | Men | 13.19 | 12.36 | 14.03 | Observed_toft |
| **Tokelau** | 2014 | Women | 7.89 | 7.45 | 8.33 | Observed_intersalt |
| **Tokelau** | 2014 | Women | 12.38 | 11.16 | 13.59 | Observed_kawasaki |
| **Tokelau** | 2014 | Women | 10.22 | 9.46 | 10.98 | Observed_tanaka |
| **Tokelau** | 2014 | Women | 8.38 | 8.07 | 8.68 | Observed_toft |
| **Turkmenistan** | 2018 | Men | 8.87 | 8.72 | 9.01 | Observed_intersalt |
| **Turkmenistan** | 2018 | Men | 12.05 | 11.85 | 12.25 | Observed_kawasaki |
| **Turkmenistan** | 2018 | Men | 8.8 | 8.69 | 8.92 | Observed_tanaka |
| **Turkmenistan** | 2018 | Men | 11.16 | 11.03 | 11.28 | Observed_toft |
| **Turkmenistan** | 2018 | Women | 6.79 | 6.7 | 6.89 | Observed_intersalt |
| **Turkmenistan** | 2018 | Women | 10.07 | 9.9 | 10.24 | Observed_kawasaki |
| **Turkmenistan** | 2018 | Women | 8.54 | 8.41 | 8.67 | Observed_tanaka |
| **Turkmenistan** | 2018 | Women | 7.77 | 7.72 | 7.82 | Observed_toft |
| **Zambia** | 2017 | Men | 8.47 | 8.15 | 8.8 | Observed_intersalt |
| **Zambia** | 2017 | Men | 12.52 | 11.85 | 13.19 | Observed_kawasaki |
| **Zambia** | 2017 | Men | 8.66 | 8.31 | 9.02 | Observed_tanaka |
| **Zambia** | 2017 | Men | 11.37 | 10.95 | 11.78 | Observed_toft |
| **Zambia** | 2017 | Women | 7.14 | 6.96 | 7.32 | Observed_intersalt |
| **Zambia** | 2017 | Women | 11.47 | 10.98 | 11.97 | Observed_kawasaki |
| **Zambia** | 2017 | Women | 9.02 | 8.71 | 9.33 | Observed_tanaka |
| **Zambia** | 2017 | Women | 8.09 | 7.95 | 8.23 | Observed_toft |

**Supplementary Table 6.** Weighted distribution of predictors in each of the 49 national surveys included in the application of the model herein developed.

| Country | Year | Region | Sample size | Mean age (years) | Age range (years) | Proportion of men (%) | Mean, minimum and maximum values of SBP (mmHg) | Mean, minimum and maximum values of DBP (mmHg) | Mean, minimum and maximum values of weight (kg) | Mean, minimum and maximum values of height (m) |
| --- | --- | --- | --- | --- | --- | --- | --- | --- | --- | --- |
| Algeria | 2017 | Africa | 6536 | 38 | 18-69 | 51.7 | 126.6 (77-227) | 75.2 (32-137) | 73.3 (25-174) | 1.67 (1.02-2.05) |
| American Samoa | 2004 | Americas | 2042 | 40 | 25-64 | 50.3 | 130.7 (84.5-230.5) | 82.4 (46.5-134.5) | 100.4 (38.6-219.1) | 1.69 (1.36-2.19) |
| Bahamas | 2012 | Americas | 1400 | 42 | 24-64 | 49.9 | 127.4 (73-248.5) | 81.6 (32.5-139.5) | 84.8 (27.9-184.9) | 1.67 (1.15-2.03) |
| Barbados | 2007 | Americas | 282 | 43 | 25-69 | 51.9 | 121.9 (86-191) | 80.1 (55-115) | 77.5 (40.6-232.1) | 1.67 (1.17-1.93) |
| Benin | 2015 | Africa | 4841 | 34 | 18-69 | 49.6 | 125.8 (74.5-254.5) | 81.6 (45-142.5) | 62.3 (30-167) | 1.64 (1.21-1.98) |
| Botswana | 2014 | Africa | 3894 | 33 | 15-69 | 52.1 | 127.5 (84.5-262.5) | 80 (47-148.5) | 63.9 (31.7-171.1) | 1.66 (1.02-2) |
| British Virgin Islands | 2009 | Americas | 1067 | 43 | 25-64 | 54.1 | 130.5 (81-225.5) | 80.3 (47.5-126) | 83.2 (39.6-176.9) | 1.7 (1.14-2.26) |
| Cabo Verde | 2007 | Africa | 1723 | 38 | 25-64 | 50.3 | 133.3 (86-233.5) | 79.7 (47.5-140) | 68.3 (35-150) | 1.68 (1.23-1.96) |
| Cambodia | 2010 | Western Pacific | 5223 | 40 | 25-64 | 49.4 | 116.2 (70.5-225.5) | 72.4 (41.5-138) | 53.7 (21.1-111) | 1.57 (1.24-1.85) |
| Cayman Islands | 2012 | Americas | 1229 | 42 | 24-64 | 50.7 | 124.7 (84-207.5) | 76 (46.5-127) | 82.3 (31-196) | 1.69 (1-2.1) |
| **Comoros** | 2011 | Africa | 5029 | 39 | 25-64 | 52.6 | 127.6 (82.5-236.5) | 78.5 (48.5-144) | 64.2 (23.5-166) | 1.61 (1-2.15) |
| **Cook Islands** | 2015 | Western Pacific | 879 | 39 | 18-64 | 46.5 | 128 (91.5-194) | 78.8 (45-118.5) | 98.6 (49.1-205.1) | 1.69 (1.07-1.96) |
| **Ecuador** | 2018 | Americas | 4466 | 40 | 18-69 | 49.4 | 119.9 (78-220.5) | 76 (42-130) | 69.2 (33.4-198.4) | 1.59 (1.24-1.93) |
| **Eritrea** | 2010 | Africa | 5651 | 42 | 25-69 | 17.2 | 117 (71.5-230.5) | 74.3 (45.5-130.5) | 51.8 (28.1-99.1) | 1.6 (1.16-1.89) |
| **Eswatini** | 2014 | Africa | 3042 | 31 | 15-69 | 47.4 | 124.3 (72-251.5) | 79.7 (42-149.5) | 67.8 (22.2-227.6) | 1.63 (1.01-2.02) |
| **Ethiopia** | 2015 | Africa | 9270 | 31 | 15-69 | 56.1 | 119.7 (71-250.5) | 77.7 (30-142) | 54.4 (20-99.5) | 1.63 (1.05-2) |
| **Fiji** | 2011 | Western Pacific | 2492 | 42 | 25-64 | 51 | 130.3 (84.5-228) | 80.2 (39-143) | 78.6 (30.3-198.1) | 1.68 (1.03-1.94) |
| **French Polynesia** | 2010 | Western Pacific | 2239 | 36 | 18-64 | 50.7 | 125 (85.5-230.5) | 79.4 (48-149.5) | 86.2 (41-193) | 1.7 (1.41-2) |
| **Gambia** | 2010 | Africa | 3496 | 38 | 25-64 | 50.4 | 130.4 (85-252) | 80.3 (44-143.5) | 64.8 (26.5-168.9) | 1.64 (1-2) |
| **Grenada** | 2011 | Americas | 1055 | 41 | 25-64 | 50.7 | 130.5 (71-212.5) | 80.5 (49.5-128) | 77.6 (40.8-158.8) | 1.7 (1.32-2.49) |
| **Guyana** | 2016 | Americas | 2625 | 37 | 18-69 | 52 | 125.7 (73.5-245) | 77.6 (37-149) | 69.9 (26.4-198) | 1.63 (1.01-2.07) |
| **Iraq** | 2015 | Eastern Mediterranean | 3655 | 35 | 18-69 | 53.6 | 128.1 (78.5-225) | 82.7 (45-150) | 76.5 (36.6-187.2) | 1.65 (1.01-1.97) |
| **Kenya** | 2015 | Africa | 4270 | 34 | 16-69 | 50.6 | 124.7 (76.5-262) | 80.8 (46.5-146) | 63.2 (30-171.3) | 1.65 (1.01-1.95) |
| **Kiribati** | 2016 | Western Pacific | 1240 | 40 | 18-69 | 42.8 | 128 (85-219.5) | 84.6 (49-147.5) | 81.1 (30-219) | 1.64 (1.22-1.89) |
| **Kuwait** | 2014 | Eastern Mediterranean | 2871 | 36 | 18-69 | 49.5 | 120.4 (70-240) | 77.4 (50-130) | 80.5 (37.3-195) | 1.65 (1.04-1.96) |
| **Kyrgyzstan** | 2013 | Eastern Mediterranean | 2539 | 41 | 25-64 | 51.9 | 133.2 (81.5-244.5) | 86.8 (56.5-149.5) | 71.7 (36.6-162.4) | 1.64 (1.38-1.95) |
| **Lao People's Democratic Republic** | 2013 | Western Pacific | 2464 | 39 | 16-65 | 42.3 | 118.7 (72-239.5) | 76.4 (30-130) | 54.2 (27-103.1) | 1.54 (1.16-1.97) |
| **Lesotho** | 2012 | Africa | 2162 | 38 | 25-64 | 49.8 | 126.3 (77.5-250) | 82.6 (46.5-146) | 66.2 (21.5-164.6) | 1.61 (1.02-1.97) |
| **Liberia** | 2011 | Africa | 2242 | 40 | 25-64 | 50.7 | 129 (87.5-232) | 79.8 (32-137.5) | 65.4 (32-163) | 1.58 (1-2.5) |
| **Libya** | 2009 | Eastern Mediterranean | 3223 | 37 | 25-64 | 51.5 | 133.4 (74-237.5) | 79.5 (44-148.5) | 77 (31.7-186.2) | 1.67 (1-1.97) |
| **Myanmar** | 2014 | Southeast Asia | 7892 | 42 | 25-64 | 50.4 | 126.1 (70-252) | 81.8 (35-144) | 57.1 (26.3-173) | 1.59 (1-2.18) |
| **Nauru** | 2016 | Western Pacific | 1037 | 36 | 18-69 | 50 | 122.6 (76-223) | 80.5 (46.5-125) | 92.4 (43.4-197.9) | 1.63 (1.41-1.86) |
| **Niue** | 2012 | Western Pacific | 779 | 40 | 15-69 | 50.1 | 127.5 (89-223) | 75.7 (44.5-117) | 91.5 (44.7-165.9) | 1.69 (1.17-1.96) |
| **Palau** | 2013 | Western Pacific | 2148 | 43 | 25-64 | 53 | 138.3 (87-236) | 84.7 (40-135) | 79.4 (32-180.6) | 1.62 (1.02-2.03) |
| **Qatar** | 2012 | Eastern Mediterranean | 2287 | 35 | 18-64 | 50.9 | 118.5 (78.5-203) | 78.8 (46.5-129.5) | 79.1 (34.4-190.5) | 1.64 (1.35-2) |
| **Republic of Moldova** | 2013 | Europe | 4077 | 39 | 18-69 | 52.5 | 132.9 (83-257) | 85.1 (49-148) | 75 (32.5-166) | 1.68 (1.2-1.98) |
| **Rwanda** | 2013 | Africa | 6882 | 32 | 15-64 | 48.8 | 120.7 (75-250) | 77.9 (45-140) | 57 (23.1-165.8) | 1.6 (1-1.91) |
| **Samoa** | 2013 | Western Pacific | 1490 | 37 | 18-64 | 54.1 | 124.9 (80.5-221.5) | 75.3 (44.5-131.5) | 90.3 (32.1-160) | 1.68 (1.22-1.97) |
| **Sao Tome and Principe** | 2008 | Africa | 2272 | 40 | 25-64 | 48.4 | 135 (77.5-240.5) | 82.5 (34.5-143) | 66.1 (30-186.2) | 1.64 (1.01-1.98) |
| **Sierra Leone** | 2009 | Africa | 4473 | 40 | 25-64 | 50.3 | 131.3 (72-220) | 80.5 (42.5-148) | 60 (28-185) | 1.62 (1-2.34) |
| **Sri Lanka** | 2015 | Southeast Asia | 4566 | 39 | 18-69 | 51.5 | 125.3 (74-258.5) | 80.6 (36-150) | 58 (26.2-156.9) | 1.59 (1.02-1.9) |
| **Tajikistan** | 2017 | Europe | 2643 | 32 | 18-69 | 53.8 | 128.6 (81-267) | 84.4 (54.5-149.5) | 66.7 (27.8-148) | 1.63 (1.09-2) |
| **Timor-Leste** | 2014 | Southeast Asia | 2480 | 36 | 18-69 | 63.8 | 130.4 (72.5-235) | 84 (42.5-136) | 52 (27-165) | 1.57 (1.24-1.83) |
| **Togo** | 2011 | Africa | 3995 | 32 | 15-64 | 49.3 | 123 (70-251) | 76.8 (31-141.5) | 61.6 (26-165) | 1.64 (1.02-1.99) |
| **Tuvalu** | 2015 | Western Pacific | 1024 | 39 | 18-69 | 54.9 | 134.4 (92.5-246) | 84.4 (48.5-145) | 91.9 (35.8-181.8) | 1.68 (1.17-2.06) |
| **Uganda** | 2014 | Africa | 3673 | 35 | 18-69 | 50.5 | 125 (83-249) | 80.8 (50-148) | 59.4 (30.2-165) | 1.62 (1.15-2.03) |
| **United Republic of Tanzania** | 2012 | Africa | 5381 | 39 | 25-64 | 50.6 | 129.1 (80-240) | 80.5 (40-145.5) | 60.6 (29-171.1) | 1.63 (1.13-1.97) |
| **Vanuatu** | 2011 | Western Pacific | 4420 | 40 | 25-64 | 47.7 | 129.6 (77-269) | 79.6 (37.5-139) | 69.4 (28.3-199.8) | 1.63 (1.02-2.1) |
| **Vietnam** | 2015 | Western Pacific | 3033 | 39 | 18-69 | 50.4 | 120 (71-223.5) | 77.2 (40-127.5) | 54.7 (27.8-106.4) | 1.58 (1.01-1.98) |

**Supplementary Table 7.** Predicted mean salt intake (g/day) by sex in each of the 49 national surveys included in the application of the model herein developed.

| Country | Year | Sex | Mean salt intake (g/day) | Mean salt intake (g/day) lower | Mean salt intake (g/day) upper | Category |
| --- | --- | --- | --- | --- | --- | --- |
| Algeria | 2017 | Men | 9.43 | 9.37 | 9.48 | predicted |
| Algeria | 2017 | Women | 7.72 | 7.68 | 7.76 | predicted |
| Algeria | 2017 | Total | 8.6 | 8.56 | 8.64 | predicted |
| American Samoa | 2004 | Men | 11.06 | 10.99 | 11.13 | predicted |
| American Samoa | 2004 | Women | 9.01 | 8.95 | 9.06 | predicted |
| American Samoa | 2004 | Total | 10.04 | 10 | 10.08 | predicted |
| Bahamas | 2012 | Men | 10.39 | 10.21 | 10.58 | predicted |
| Bahamas | 2012 | Women | 8.26 | 8.01 | 8.51 | predicted |
| Bahamas | 2012 | Total | 9.33 | 9.19 | 9.46 | predicted |
| Barbados | 2007 | Men | 9.61 | 9.31 | 9.92 | predicted |
| Barbados | 2007 | Women | 7.86 | 7.54 | 8.18 | predicted |
| Barbados | 2007 | Total | 8.77 | 8.56 | 8.98 | predicted |
| Benin | 2015 | Men | 8.81 | 8.72 | 8.91 | predicted |
| Benin | 2015 | Women | 7.29 | 7.2 | 7.37 | predicted |
| Benin | 2015 | Total | 8.04 | 7.92 | 8.16 | predicted |
| Botswana | 2014 | Men | 8.61 | 8.54 | 8.69 | predicted |
| Botswana | 2014 | Women | 7.39 | 7.34 | 7.45 | predicted |
| Botswana | 2014 | Total | 8.03 | 7.97 | 8.09 | predicted |
| British Virgin Islands | 2009 | Men | 10.09 | 10.03 | 10.16 | predicted |
| British Virgin Islands | 2009 | Women | 8.02 | 8.01 | 8.02 | predicted |
| British Virgin Islands | 2009 | Total | 9.14 | 9.08 | 9.2 | predicted |
| Cabo Verde | 2007 | Men | 9.13 | 9 | 9.25 | predicted |
| Cabo Verde | 2007 | Women | 7.46 | 7.36 | 7.55 | predicted |
| Cabo Verde | 2007 | Total | 8.3 | 8.19 | 8.41 | predicted |
| Cambodia | 2010 | Men | 8.7 | 8.66 | 8.75 | predicted |
| Cambodia | 2010 | Women | 7.23 | 7.2 | 7.25 | predicted |
| Cambodia | 2010 | Total | 7.96 | 7.92 | 7.99 | predicted |
| Cayman Islands | 2012 | Men | 9.95 | 9.89 | 10 | predicted |
| Cayman Islands | 2012 | Women | 8.08 | 7.86 | 8.29 | predicted |
| Cayman Islands | 2012 | Total | 9.03 | 8.96 | 9.09 | predicted |
| Comoros | 2011 | Men | 9.09 | 9.04 | 9.15 | predicted |
| **Comoros** | 2011 | Women | 7.74 | 7.7 | 7.78 | predicted |
| **Comoros** | 2011 | Total | 8.45 | 8.4 | 8.5 | predicted |
| **Cook Islands** | 2015 | Men | 11.02 | 10.95 | 11.1 | predicted |
| **Cook Islands** | 2015 | Women | 8.63 | 8.56 | 8.69 | predicted |
| **Cook Islands** | 2015 | Total | 9.74 | 9.64 | 9.84 | predicted |
| **Ecuador** | 2018 | Men | 9.74 | 9.67 | 9.81 | predicted |
| **Ecuador** | 2018 | Women | 7.8 | 7.75 | 7.84 | predicted |
| **Ecuador** | 2018 | Total | 8.76 | 8.68 | 8.83 | predicted |
| **Eritrea** | 2010 | Men | 8.4 | 8.34 | 8.47 | predicted |
| **Eritrea** | 2010 | Women | 6.93 | 6.89 | 6.97 | predicted |
| **Eritrea** | 2010 | Total | 7.18 | 7.14 | 7.22 | predicted |
| **Eswatini** | 2014 | Men | 9.01 | 8.9 | 9.12 | predicted |
| **Eswatini** | 2014 | Women | 7.71 | 7.65 | 7.77 | predicted |
| **Eswatini** | 2014 | Total | 8.33 | 8.26 | 8.39 | predicted |
| **Ethiopia** | 2015 | Men | 8.28 | 8.24 | 8.32 | predicted |
| **Ethiopia** | 2015 | Women | 6.92 | 6.89 | 6.95 | predicted |
| **Ethiopia** | 2015 | Total | 7.69 | 7.65 | 7.72 | predicted |
| **Fiji** | 2011 | Men | 9.75 | 9.63 | 9.86 | predicted |
| **Fiji** | 2011 | Women | 8.02 | 7.95 | 8.08 | predicted |
| **Fiji** | 2011 | Total | 8.9 | 8.79 | 9.01 | predicted |
| **French Polynesia** | 2010 | Men | 10.24 | 10.13 | 10.35 | predicted |
| **French Polynesia** | 2010 | Women | 8.04 | 7.95 | 8.13 | predicted |
| **French Polynesia** | 2010 | Total | 9.16 | 9.06 | 9.25 | predicted |
| **Gambia** | 2010 | Men | 9.1 | 8.96 | 9.24 | predicted |
| **Gambia** | 2010 | Women | 7.54 | 7.46 | 7.62 | predicted |
| **Gambia** | 2010 | Total | 8.33 | 8.22 | 8.43 | predicted |
| **Grenada** | 2011 | Men | 9.49 | 9.39 | 9.59 | predicted |
| **Grenada** | 2011 | Women | 7.89 | 7.8 | 7.98 | predicted |
| **Grenada** | 2011 | Total | 8.7 | 8.61 | 8.79 | predicted |
| **Guyana** | 2016 | Men | 9.28 | 9.17 | 9.39 | predicted |
| **Guyana** | 2016 | Women | 7.85 | 7.78 | 7.92 | predicted |
| **Guyana** | 2016 | Total | 8.59 | 8.53 | 8.66 | predicted |
| **Iraq** | 2015 | Men | 9.73 | 9.63 | 9.84 | predicted |
| **Iraq** | 2015 | Women | 8.01 | 7.95 | 8.07 | predicted |
| **Iraq** | 2015 | Total | 8.94 | 8.86 | 9.01 | predicted |
| **Kenya** | 2015 | Men | 8.69 | 8.59 | 8.78 | predicted |
| **Kenya** | 2015 | Women | 7.35 | 7.27 | 7.42 | predicted |
| **Kenya** | 2015 | Total | 8.02 | 7.94 | 8.11 | predicted |
| **Kiribati** | 2016 | Men | 10.19 | 9.99 | 10.39 | predicted |
| **Kiribati** | 2016 | Women | 8.25 | 8.14 | 8.36 | predicted |
| **Kiribati** | 2016 | Total | 9.08 | 8.94 | 9.23 | predicted |
| **Kuwait** | 2014 | Men | 10.19 | 10.11 | 10.26 | predicted |
| **Kuwait** | 2014 | Women | 8.08 | 8.03 | 8.13 | predicted |
| **Kuwait** | 2014 | Total | 9.13 | 9.07 | 9.18 | predicted |
| **Kyrgyzstan** | 2013 | Men | 9.58 | 9.43 | 9.73 | predicted |
| **Kyrgyzstan** | 2013 | Women | 7.8 | 7.75 | 7.85 | predicted |
| **Kyrgyzstan** | 2013 | Total | 8.72 | 8.65 | 8.8 | predicted |
| **Lao People's Democratic Republic** | 2013 | Men | 8.97 | 8.88 | 9.05 | predicted |
| **Lao People's Democratic Republic** | 2013 | Women | 7.38 | 7.32 | 7.43 | predicted |
| **Lao People's Democratic Republic** | 2013 | Total | 8.05 | 7.98 | 8.12 | predicted |
| **Lesotho** | 2012 | Men | 9.01 | 8.9 | 9.11 | predicted |
| **Lesotho** | 2012 | Women | 7.92 | 7.83 | 8 | predicted |
| **Lesotho** | 2012 | Total | 8.46 | 8.38 | 8.54 | predicted |
| **Liberia** | 2011 | Men | 9.52 | 9.38 | 9.65 | predicted |
| **Liberia** | 2011 | Women | 7.93 | 7.82 | 8.05 | predicted |
| **Liberia** | 2011 | Total | 8.74 | 8.61 | 8.87 | predicted |
| **Libya** | 2009 | Men | 9.67 | 9.59 | 9.75 | predicted |
| **Libya** | 2009 | Women | 8.07 | 8 | 8.14 | predicted |
| **Libya** | 2009 | Total | 8.89 | 8.83 | 8.95 | predicted |
| **Myanmar** | 2014 | Men | 8.71 | 8.59 | 8.82 | predicted |
| **Myanmar** | 2014 | Women | 7.41 | 7.32 | 7.49 | predicted |
| **Myanmar** | 2014 | Total | 8.06 | 7.98 | 8.14 | predicted |
| **Nauru** | 2016 | Men | 11.11 | 11.01 | 11.2 | predicted |
| **Nauru** | 2016 | Women | 8.76 | 8.6 | 8.92 | predicted |
| **Nauru** | 2016 | Total | 9.93 | 9.8 | 10.06 | predicted |
| **Niue** | 2012 | Men | 10.73 | 10.61 | 10.85 | predicted |
| **Niue** | 2012 | Women | 8.33 | 8.21 | 8.44 | predicted |
| **Niue** | 2012 | Total | 9.53 | 9.41 | 9.65 | predicted |
| **Palau** | 2013 | Men | 10.52 | 10.4 | 10.65 | predicted |
| **Palau** | 2013 | Women | 8.19 | 8.12 | 8.26 | predicted |
| **Palau** | 2013 | Total | 9.43 | 9.33 | 9.53 | predicted |
| **Qatar** | 2012 | Men | 10.17 | 10.06 | 10.29 | predicted |
| **Qatar** | 2012 | Women | 8.11 | 8.01 | 8.21 | predicted |
| **Qatar** | 2012 | Total | 9.16 | 9.06 | 9.26 | predicted |
| **Republic of Moldova** | 2013 | Men | 9.64 | 9.56 | 9.72 | predicted |
| **Republic of Moldova** | 2013 | Women | 7.56 | 7.5 | 7.62 | predicted |
| **Republic of Moldova** | 2013 | Total | 8.65 | 8.59 | 8.72 | predicted |
| **Rwanda** | 2013 | Men | 8.67 | 8.63 | 8.7 | predicted |
| **Rwanda** | 2013 | Women | 7.29 | 7.26 | 7.32 | predicted |
| **Rwanda** | 2013 | Total | 7.96 | 7.93 | 7.99 | predicted |
| **Samoa** | 2013 | Men | 10.44 | 10.3 | 10.58 | predicted |
| **Samoa** | 2013 | Women | 8.63 | 8.56 | 8.7 | predicted |
| **Samoa** | 2013 | Total | 9.61 | 9.51 | 9.7 | predicted |
| **Sao Tome and Principe** | 2008 | Men | 9.11 | 9.07 | 9.15 | predicted |
| **Sao Tome and Principe** | 2008 | Women | 7.54 | 7.44 | 7.64 | predicted |
| **Sao Tome and Principe** | 2008 | Total | 8.3 | 8.22 | 8.38 | predicted |
| **Sierra Leone** | 2009 | Men | 8.83 | 8.73 | 8.93 | predicted |
| **Sierra Leone** | 2009 | Women | 7.41 | 7.32 | 7.51 | predicted |
| **Sierra Leone** | 2009 | Total | 8.13 | 8.03 | 8.23 | predicted |
| **Sri Lanka** | 2015 | Men | 8.82 | 8.75 | 8.89 | predicted |
| **Sri Lanka** | 2015 | Women | 7.29 | 7.25 | 7.33 | predicted |
| **Sri Lanka** | 2015 | Total | 8.08 | 8.03 | 8.13 | predicted |
| **Tajikistan** | 2017 | Men | 9.4 | 9.3 | 9.5 | predicted |
| **Tajikistan** | 2017 | Women | 7.55 | 7.48 | 7.62 | predicted |
| **Tajikistan** | 2017 | Total | 8.55 | 8.46 | 8.64 | predicted |
| **Timor-Leste** | 2014 | Men | 8.42 | 8.37 | 8.46 | predicted |
| **Timor-Leste** | 2014 | Women | 7 | 6.88 | 7.11 | predicted |
| **Timor-Leste** | 2014 | Total | 7.9 | 7.8 | 8 | predicted |
| **Togo** | 2011 | Men | 8.71 | 8.66 | 8.77 | predicted |
| **Togo** | 2011 | Women | 7.21 | 7.16 | 7.26 | predicted |
| **Togo** | 2011 | Total | 7.95 | 7.9 | 8 | predicted |
| **Tuvalu** | 2015 | Men | 10.57 | 10.44 | 10.71 | predicted |
| **Tuvalu** | 2015 | Women | 8.59 | 8.5 | 8.68 | predicted |
| **Tuvalu** | 2015 | Total | 9.68 | 9.57 | 9.79 | predicted |
| **Uganda** | 2014 | Men | 8.63 | 8.57 | 8.69 | predicted |
| **Uganda** | 2014 | Women | 7.22 | 7.16 | 7.28 | predicted |
| **Uganda** | 2014 | Total | 7.93 | 7.87 | 7.99 | predicted |
| **United Republic of Tanzania** | 2012 | Men | 8.6 | 8.47 | 8.72 | predicted |
| **United Republic of Tanzania** | 2012 | Women | 7.48 | 7.41 | 7.55 | predicted |
| **United Republic of Tanzania** | 2012 | Total | 8.04 | 7.98 | 8.11 | predicted |
| **Vanuatu** | 2011 | Men | 9.52 | 9.44 | 9.59 | predicted |
| **Vanuatu** | 2011 | Women | 7.75 | 7.69 | 7.81 | predicted |
| **Vanuatu** | 2011 | Total | 8.59 | 8.53 | 8.66 | predicted |
| **Vietnam** | 2015 | Men | 8.75 | 8.68 | 8.82 | predicted |
| **Vietnam** | 2015 | Women | 7.11 | 7.07 | 7.16 | predicted |
| **Vietnam** | 2015 | Total | 7.94 | 7.88 | 8 | predicted |

**Supplementary Table 8.** Comparison between mean salt intake predictions and global estimates across national surveys included in the ML application.

| Country | Year (for ML predictions) | ML-predicted mean salt intake and 95% CI (g/day) | Year (for global estimates) | Estimated mean salt intake and 95% CI (g/day) | Ratio between predicted and estimated mean salt intake | Absolute difference between predicted mean salt intake and global estimated mean salt intake |
| --- | --- | --- | --- | --- | --- | --- |
| Algeria | 2017 | 8.6 (8.6-8.6) | 2010 | 10.7 (9-12.5) | 0.8 | 2.1 |
| Bahamas | 2012 | 9.3 (9.2-9.5) | 2010 | 7.5 (6.2-8.8) | 1.2 | 1.8 |
| Barbados | 2007 | 8.8 (8.6-9) | 2010 | 8.6 (7.8-9.4) | 1 | 0.2 |
| Benin | 2015 | 8 (7.9-8.2) | 2010 | 7.1 (6.2-8.1) | 1.1 | 0.9 |
| Botswana | 2014 | 8 (8-8.1) | 2010 | 6.3 (5.4-7.4) | 1.3 | 1.7 |
| Cabo Verde | 2007 | 8.3 (8.2-8.4) | 2010 | 8.1 (6.8-9.7) | 1 | 0.2 |
| Cambodia | 2010 | 8 (7.9-8) | 2010 | 11 (9.3-12.9) | 0.7 | 3 |
| Comoros | 2011 | 8.4 (8.4-8.5) | 2010 | 4.2 (3.5-5) | 2 | 4.2 |
| Ecuador | 2018 | 8.8 (8.7-8.8) | 2010 | 7.6 (6.4-8.9) | 1.2 | 1.2 |
| Eritrea | 2010 | 7.2 (7.1-7.2) | 2010 | 5.9 (5-7) | 1.2 | 1.3 |
| Ethiopia | 2015 | 7.7 (7.7-7.7) | 2010 | 5.7 (4.9-6.7) | 1.4 | 2 |
| Fiji | 2011 | 8.9 (8.8-9) | 2010 | 7.2 (6-8.5) | 1.2 | 1.7 |
| Gambia | 2010 | 8.3 (8.2-8.4) | 2010 | 7.7 (6.5-8.9) | 1.1 | 0.6 |
| Grenada | 2011 | 8.7 (8.6-8.8) | 2010 | 6.5 (5.5-7.7) | 1.3 | 2.2 |
| Guyana | 2016 | 8.6 (8.5-8.7) | 2010 | 6.1 (5.1-7.3) | 1.4 | 2.5 |
| Iraq | 2015 | 8.9 (8.9-9) | 2010 | 9.4 (8-11.2) | 0.9 | 0.5 |
| Kenya | 2015 | 8 (7.9-8.1) | 2010 | 3.7 (3.4-4) | 2.2 | 4.3 |
| Kiribati | 2016 | 9.1 (8.9-9.2) | 2010 | 5.6 (4.6-6.7) | 1.6 | 3.5 |
| Kuwait | 2014 | 9.1 (9.1-9.2) | 2010 | 9.7 (8.7-10.8) | 0.9 | 0.6 |
| Kyrgyzstan | 2013 | 8.7 (8.7-8.8) | 2010 | 13.4 (11.4-15.8) | 0.6 | 4.7 |
| **Lao People's Democratic Republic** | 2013 | 8.1 (8-8.1) | 2010 | 11.1 (9.4-13.2) | 0.7 | 3 |
| **Lesotho** | 2012 | 8.5 (8.4-8.5) | 2010 | 6.6 (5.5-7.8) | 1.3 | 1.9 |
| **Liberia** | 2011 | 8.7 (8.6-8.9) | 2010 | 6.7 (5.6-7.9) | 1.3 | 2 |
| **Libya** | 2009 | 8.9 (8.8-8.9) | 2010 | 10.6 (8.9-12.5) | 0.8 | 1.7 |
| **Myanmar** | 2014 | 8.1 (8-8.1) | 2010 | 11.2 (9.4-13.2) | 0.7 | 3.1 |
| **Qatar** | 2012 | 9.2 (9.1-9.3) | 2010 | 10.5 (8.3-12.9) | 0.9 | 1.3 |
| **Republic of Moldova** | 2013 | 8.7 (8.6-8.7) | 2010 | 9.9 (8.3-11.6) | 0.9 | 1.2 |
| **Rwanda** | 2013 | 8 (7.9-8) | 2010 | 4 (3.3-4.9) | 2 | 4 |
| **Samoa** | 2013 | 9.6 (9.5-9.7) | 2010 | 5.2 (4.6-5.8) | 1.8 | 4.4 |
| **Sao Tome and Principe** | 2008 | 8.3 (8.2-8.4) | 2010 | 5.9 (4.9-6.9) | 1.4 | 2.4 |
| **Sierra Leone** | 2009 | 8.1 (8-8.2) | 2010 | 6.3 (5.3-7.3) | 1.3 | 1.8 |
| **Sri Lanka** | 2015 | 8.1 (8-8.1) | 2010 | 9.7 (8.2-11.3) | 0.8 | 1.6 |
| **Tajikistan** | 2017 | 8.6 (8.5-8.6) | 2010 | 13.5 (11.6-15.7) | 0.6 | 4.9 |
| **Timor-Leste** | 2014 | 7.9 (7.8-8) | 2010 | 11.2 (9.3-13.3) | 0.7 | 3.3 |
| **Uganda** | 2014 | 7.9 (7.9-8) | 2010 | 5.3 (4.4-6.3) | 1.5 | 2.6 |
| **United Republic of Tanzania** | 2012 | 8 (8-8.1) | 2010 | 6.9 (6.1-7.7) | 1.2 | 1.1 |
| **Vanuatu** | 2011 | 8.6 (8.5-8.7) | 2010 | 5.6 (4.8-6.6) | 1.5 | 3 |
| **Vietnam** | 2015 | 7.9 (7.9-8) | 2010 | 11.5 (9.5-13.7) | 0.7 | 3.6 |

\*There are 38 countries in this table; that is, countries included in our analysis that were not available in the previous global work, were not included in this table.

## Notes

### Competing Interest Statement

The authors have declared no competing interest.

### Funding Statement

RMC-L is supported by a Wellcome Trust International Training Fellowship (214185/Z/18/Z).

